# Analysis of SIR-Network Model on COVID-19 with respect to its impact on West Bengal in India

**DOI:** 10.1101/2020.08.05.20169037

**Authors:** Debnarayan Khatua, Debashree Guha, Samarjit Kar, Anupam De, Eshan Samanta

**Author notes:** Email addresses* (Debnarayan Khatua), (Debashree Guha), (Samarjit Kar), (Anupam De), (Eshan Samanta).

## Abstract

The recent global pandemic of SARS-CoV-2 (COVID-19) disease has prompted many researchers to formulate lock-down and quarantine scenarios while the main concern of the researchers is to model the spread and the possible duration of the COVID-19 infections and also research on how long this is going to last. It seems that most of the researchers have recognized lock-down as one of the major impact factors in their models. As a result of this in the absence of lock-down the models formulated would not contribute much significant results. Hence, in this work we decide to formulate a mathematical model which would be able to predict the spread and also the possible duration of the pandemic, by considering both partial lock-down and the corresponding unlocking situations. Employing SIR-network models and taking the various districts of highly populated areas of West Bengal, India as the nodes or vertices we attempt to model the spread and duration of the pandemic during both partial lock-down and unlock phases but separately. We consider the populations where the locally present people and the people who have undergone migration of some shorts are well mixed together. In the network that we have provided the pointed edges refer to the migrating workers that is those that move away from their regular habitats in want of work. We use this research to study not only the trends that are associated with COVID-19 outbreaks, but also to study the impacts of the Government policies and the improvisation of medical facilities on this outbreak in West Bengal. At the end, we attempt to throw light upon the crisis that the economy of the state may have to go through separately on partial lock-down and unlocking scenarios.

## 1. Introduction

In December 2019, an outbreak of pneumonia of unknown cause was originated, from Wuhan city, the capital of Hubei province in China [1,2], and quickly spread to all Chinese provinces. Despite of introducing prompt radical measures to control the outbreak [3], over the course of less than three months it has spread to 58 other countries [4] and was diagnosed as corona virus disease 2019 (COVID-19) in February, 2020, by WHO. The World Health Organisation report dated 25th May 2020 reported 53,04,772 total cases and 3,42,029 deaths worldwide [5].

India has been severely affected. After the first indigenous case on 30th January 2020 in Kerala (southern province) [6], very few new cases were discovered during February. Over the month of March, several cases began to emerge across India [7, 8, 9] and India enters the World Health Organization’s (WHO) list of areas where the virus has been reported. As a consequence, multiple states across the country began shutting down schools, colleges, public facilities such as malls, gyms, cinema halls and other public places to contain the spread [10, 11]. Further, due to the mass spread of infection, and increasing pressure on hospital capacity following the examples of other countries India officially implemented: lock-downs, mobility restrictions, quarantine hospitals, increase in public health measures, protection of the elderly, etc. [12]. As per Ministry of Home Affairs (MHA) order [13] a district-wise zonal classification system was instituted on 3rd May 2020, comprises of a ‘red zone’, encompassing 170 hot-spot districts where SARS-CoV-2 infection was endemic, ‘green zone’ including 219 districts with zero confirmed cases till date or no confirmed case in the last 21 days, ‘orange zones’ comprising of 284 districts which are neither in the Red zone nor in the Green zone, and put on the extension of the second phase of the nationwide lock-down to contain the emerging threat. Although different corrective measures have slowed the spread of infection, the cost to the economy of such measures is considerable. For instance, with the first 1.5 months of lock-down in India it is estimated that the crisis is expected to cost nearly 4percentage of its GDP [14]. Beginning 1st June the Government has started unlocking the country (barring containment zones) in three unlock phases.

COVID-19 is an acute and new disease spread by a virus belongs to corona family that is growing significantly worldwide with a name known as SARS-CoV-2 and are related to genetically the one that caused the outbreak of Severe Acute Respiratory Syndrome (SARS) in 2003, but the diseases they cause are quite different. SARS was more deadly but much less infectious than COVID-19. The estimated basic reproductive values (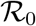) of SARS-CoV-2 at the early stage were calculated between 2 and 3.5, indicating that one patient could transmit the disease to two to three other people, [15, 16, 17] which was higher than SARS and MERS. A study from China showed [18], a large number of transmissions can be occurred, both in nosocomial and community settings. Through feces, and aerosols transmission SARS-CoV-2 spread is also highly possible. The viral load of SARS-CoV-2 detected in the asymptomatic patients was similar to that in the symptomatic patients. Majority of infected individuals with no or mild symptoms can release viruses while coughs or exhales and spread viruses to others through small droplets, and becomes the cause of pandemic disease associated with substantial morbidity and mortality within a period of weeks, which is extremely challenging for preventing the spread of COVID-19 without the exploration of a vaccine. Viral shedding pattern of the patients infected with SARS-COV-2 is analogous to that of influenza viruses [19]. These findings suggest that, in-apparent transmission may play an important role in sustaining the outbreak.

To understand the process of disease transmission during the course of the epidemic and how to plan effective control strategies Predictive mathematical models have been studied for decades [20, 21, 22, 23]. One commonly used compartmental model is the SIR model [24] for portraying the dynamics of infectious disease transmission, which describes the flow of individuals with three mutually exclusive stages of infection or compartments: susceptible, infected and recovered. More complex models can be utilized for modeling infectious disease dynamics. Researchers [25, 26, 27, 28] have developed several mathematical models for COVID 19 pandemic. As well as this, Lin and colleagues [25] have developed an SIR model considering risk perception and the number of cases, Anastassopoulou et al. [26] have proposed a separate time SIR model with the dead populations, Casella [27] has developed a control-based SIR model that emphasizes the effects of delays and compares results on the basis of containment policies, Wu and colleagues [28] have used infection dynamics to estimate the clinical severity of COVID-19. Wang et al.[29] have presented some critical reviews for mathematical models for the prevalence of COVID-19. Fang et al. [30] have presented an overall analysis of the transmission rate due to COVID-19 using the SEIR model. Hou et al. [31] have reported the quarantine effectiveness against coronavirus 2019 (COVD-19) in Wuhan, China. Radulescu and Cavanagh [32] have discussed the management strategies of COVID 19 community spreads using the SEIR model. Pandey et al. [33] have predicted the outbreak of COVID-19 in India based on SEIR and regression models. Stochastic transmission models have also been considered [34, 35]. Besides that, Ndairou et al.[36] have proposed a sectional mathematical model based on SEIPAHRF for the proliferation of COVID-19 disease with a special focus on the contagiousness of super-spreaders. Recently, Khatua et al. [37] have predicted the pandemic on Indian aspect by control-based SEIAR model and also have developed a control-based SEIAHRD model [38] with the uncertain transmission parameters.

Here we propose to employ a SIR-network model for the COVID-19 epidemic in India, that extends the classical SIR model, to capture the impact of human mobility in future progress of the disease. SIR-network model was introduced in [39, 40, 41, 42] for analyzing the epidemic models into networks. These models consider that the ideal networks are made up of nodes that represent regions/ affected regions/affected areas and links that mimic interactions or connections between them. In our study we consider SIR-network model as a Predictive model with the aims to (i)understand the consequence of unlocking decision in the future disease progression (ii) recognize the facts that the infection with COVID-19 may reach its saturation level within a time frame after an easing of partial lockdown restrictions provided social distancing is maintained. Our model does not consider reduced availability of medical heath care i.e., recovery rate of the infection is retained trough out the simulation. Specifically, in designing this model as a simplification of COVID-19 dynamics, local transmission parameters reveal variations in environmental characteristics such as recovery rate, death rate, transmission rate. Here we stress on a heterogeneous network to explore the mobility of people to be an essential ingredient in analyzing the epidemic. The focus of our analysis is to understand the COVID-19 epidemic outbreak pattern during and after partial lock-down restrictions by which to find the pathway of releasing the people from an uncertain black box and enable then to plan the way forward.

## 2. The model formulation

In this part, we have pulled in some discussions about the fundamentals of a meta-population model [43, 44, 45, 46, 42]. Metaphorical models are usually applied to identify the kinetics of an epidemic in various environment. For example, Arino and van den Driessche [45] have analyzed the spread of a disease among different cities and have thought the possibility of migration between them. Their findings explore the importance of human mobility in epidemic problems. Nevertheless, in the case of COVID-19 in an undivided province, we are interested in daily movement between neighborhoods. This is why the short-term scale of our model is one day, and we can put aside the tedious rate of migration and demographics. The idea of thinking to build up a metaphorical network is interesting. We can imagine a large state being roughly divided into districts with a population of millions. The choice of this scale should be driven by the number of COVID-19 patients and the available detailed information on the human travel pattern between the intermediate districts. The effect of the geometry of human transport networks on the spread of disease is another interesting aspect to be explored. This can be measured using road traffic studies and data from public transport companies. For a large province, it is common to get that some territories have stronger links than others.

Mathematically, we presume that the district has *X* neighborhoods (corresponding to nodes of the network) and we denote the set as *V =* (1,2,…, *n*). A node *i* is linked to another *j* if there is a fraction of residents of *i* that travel to *j*. Here we assume that the network is asymmetric means the edges are not bidirectional. In this case, the geometry of the network is then defined by specifying the flux matrix *Ψ*,

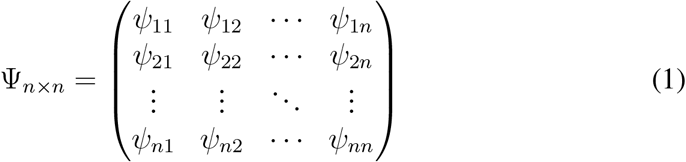

whose entries *ψ_ij_* ∈ [0, 1] are the fraction of resident population going from *i* to *j*. It is important to note that the *ψ_ij_*’s are dimensionless. As discussed in more detail [39], we assume that each node contains a conservation of the residential population, and as consequence the fraction satisfies

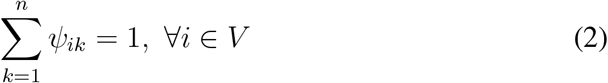

### Definition 1

*We define a homogeneous network model as one where all nodes have the same rate of transmission, η_j_* = *η*_0_, ∀*_j_* =1, 2,*…,n. All other examples are defined to be heterogeneous*.

### 2.1. SIR-Network model

A SIR model is introduced by placing individuals into three segments such as susceptible (S), infected (I) and recovered or removed (R) for the pandemic COVID-19. Susceptible is a group of people who are at risk of contracting the disease form infected people. They might become patient if infected. The infected people can pass the disease on to susceptible people and recover within a certain period of time. A susceptible person can be tainted by the virus due to close contact with an infected person. Since the disease can be transmitted from person to person, usually after close contact with an infected patient, so we take *η* here as the rate of infection or contact between susceptible or infected people. Then the part 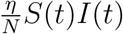 goes to the susceptible class to the infected class. We take the *δ* as the natural mortality rate per capita for each of the three segments, treatment recovery rate as *τ*, average duration of infectivity as *β* and the disease related mortality as *γ*. In our model, new births and migration are ignored. Therefore, network models with infectious energy follow dormant and infectious periods which are represented as follows

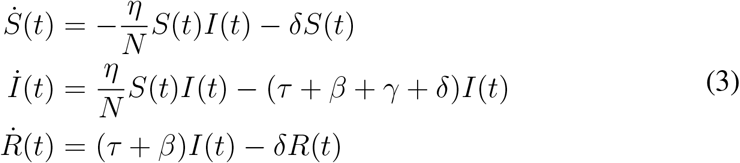

where *N* = *S* + *I* + *R* is the total population. Initially the mathematical role of epidemic problems was accomplished with such conditions *S*(0) *>* 0,*I*(0) *>* 0,*R*(0) *>* 0.

For any node *i* ∈ *V*, let *S_i_*(*t*),*I_i_*(*t*) and *R_i_*(*t*) be the number of susceptible, infectious and recovered/removed persons, respectively, in neighbourhood *i* at the time *t*. The total population individuals at *i* is considered as a constant at time *t* and is given by

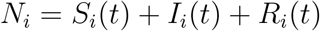

Furthermore, since *ψ_ki_N_k_* is the total population of the node *k* that travels to the node *i* every day, we can define

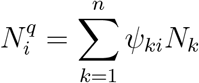

as the present population at *i*, which is also constant at the time and 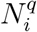 is a normalizing factor[45].

The transmission rate (or incidence rate) of an infection is the probability or risk of infection in the population. It is employed to evaluate the absorption of fresh instances of infection in a population over a point of time. For this pandemic case, we set *η_j_* as the transmission rate at the node j for all *j* ∈ *V*. For every group of populations, we follow the assumptions that the recovery rate (*τ*), the average viremic period (*β*), the disease related mortality rate (*γ*) and the natural death rate (*δ*) of every infected individuals are constants. Then the dynamical system-(3) can be expressed as the following first-order homogeneous system of differential equations

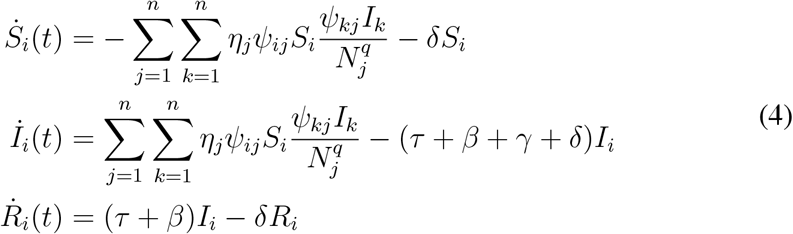

The double-summation terms enable us to model the fact that any resident of location *k* can acquire an infection at location *j*, by an individual person from location *i*, provided that the corresponding fraction *ψ_ij_* are non zero.

### 2.2. Basic reproduction number

The number of basic reproductions 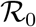 is the number of secondary cases that occur in a case of completely susceptible population. It has become common practice in the analysis of the simplest models to consider the next generation matrix process [47] and to define the basic reproduction number 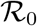 as the expected number of secondary infection produced in a fully susceptible population by a common infected person during the entire period of infection. Thus 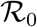 may vary significantly for different infectious diseases as well as for the same disease in different populations. The key threshold criterion then states: the disease can pandemic if 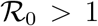, whereas it cannot if 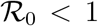. To establish the 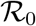 for the SIR-network model, we begin by introducing vector notation, since infected individuals are from different nodes of the network. Let 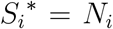 and 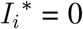 are balanced population of the susceptible and infected populations at the node *i*. We define the following notations at the time *t* as

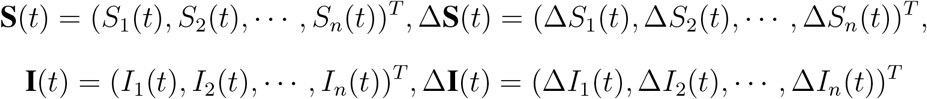

where 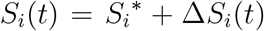 and 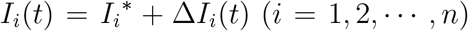 are the number of susceptible and infected persons at the vertex *i* at the time *t*. In fact, *η_ij_* depends on individual behavior, which determines the amount of mix between different groups. Replacing the outline forms in (4) and linearizing, we find

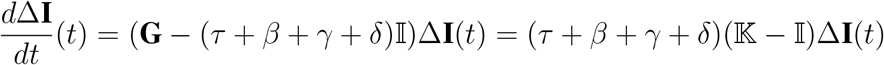

where

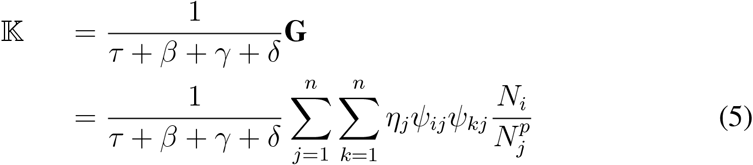

is the well-known next generation matrix(NGM) (see [48]).

#### Proposition 1

*[42] Let* (*V, ψ_n_*_×_*_n_*) *be the network given by a vertices set V* = 1, 2, ···, *n and a flux matrix ψ* =[*ψ_ij_*]*_n_*_×_*_n_ of the system (4), describing the SIR-network dynamics. Then, by considering* 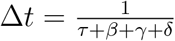 *and* 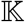 *as in (5), we have that* 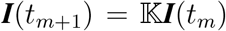 *describes the discrete dynamics of the system linearized about the equilibrium state* 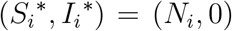 *for all i* ∈ *V. In particular, the first generation of infected individuals is given by* 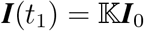.

Following the results of Proposition-1, if we identify an infected person in a disease-free population through the surrounding area *i*, *i.e*., if *I_i_*(0) = 1 for some *i* ∈ *V* and *I_j_*(0) = 0 otherwise, then the sum

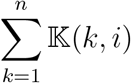

could be a good definition for 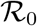. However it depends on the vertex *i*, so it can vary from one end to the other. Here we see a correspondence with the expression for the basic reproduction number of the SIR model-(3), is given by 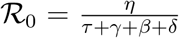. Thus, the sum 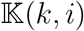 represents the average number of infected people living in vertex *k* who were infected by a single infected person introduced at vertex *i*.

#### Definition 2

*[42] Given the NGM* 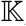, *(5), the basic reproduction number is defined to be*

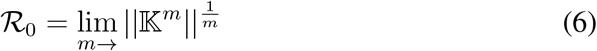

, *where* 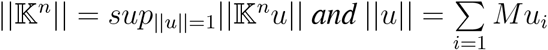.

#### Theorem 1

*Let* (*S, Ψ_n_*_×_*_n_*) *be a network associated with the SIR-network system. Let η*_0_ *>* 0 *be a constant value such that η_j_* = *η*_0_, ∀*_j_* ∈ *S. Then we have* 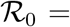 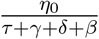, *hence epidemics are possible if and only if η*_0_ *>τ* + *γ* + *δ* + *β*.

## 3. Case study

### 3.1. Introduction

In this section we construct a simple application of SIR-Network model on the West Bengal, which is an eastern regional state of India on the Bay of Bengal. With a population of over 91 million (as of 2011), it is the fourth most populous state in India. West Bengal is also the seventh most populous regional entity in the universe. West Bengal is the thirteenth largest state in India with an expanse of 88,752 km_2_ (34,267 sq mile). It is bounded by Bangladesh to the east and Nepal and Bhutan to the northward. It is also surrounded by the Indian provinces of Orissa, Jharkhand, Bihar, Sikkim and Assam. The state capital and biggest city of the state is a Kolkata—the third-biggest urban agglomeration and the seventh-biggest metropolis in India [49]. As of 2017, West Bengal is divided into 23 districts [49] (Figure-1).

**Figure 1:**
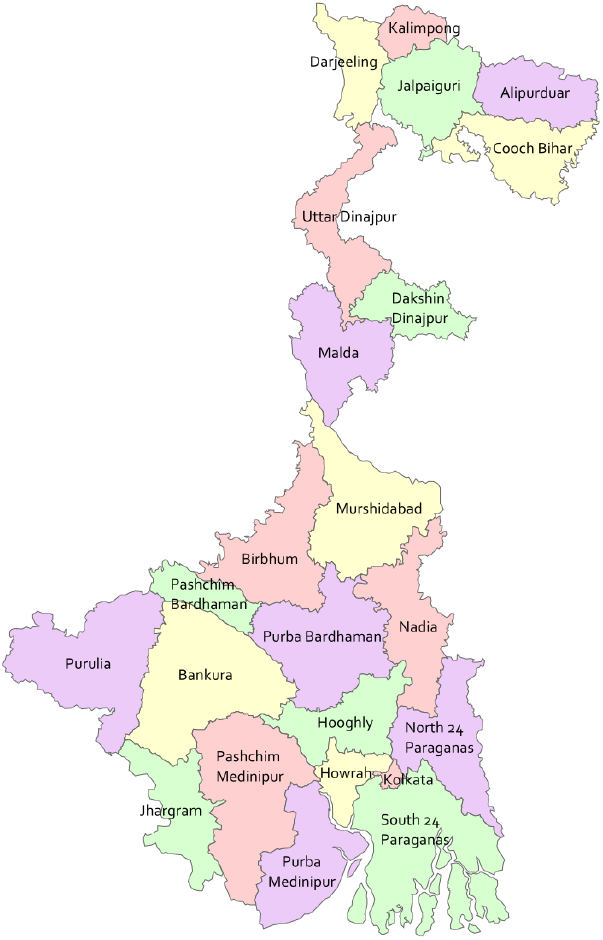
Regional map of West Bengal. We have built our small network around

Specifically, we chose to test a period when transportation between the districts has barely been started. At this time there were 3816 total registered cases and 278 total deaths in the whole District(as on 25th May,2020)[50]. In order to simplify our analysis, first task is to choose a representative part of the state. For this, instead of considering the whole of West Bengal in the simulation, we study a small but interesting regions of the whole integrated network. The network is developed by considering 8 districts connected by railways and passenger vehicle transportation system with the center Kolkata. All the selected regions are announced as a red zone or orange zone by state or central government (Figure-2). The set of regions are described in subsection-3.2. For the selected region we create a specific flux matrix(see-1 for the definition) inspired by the features that we want to find out in the network. This is made in subsection-3.3. Then, in subsection-3.4 we estimate the epidemiological parameters so that we can simplify our analysis. In subsection-3.5 we identify our assumptions regarding the data and the duration for which the fits will run.

**Figure 2:**
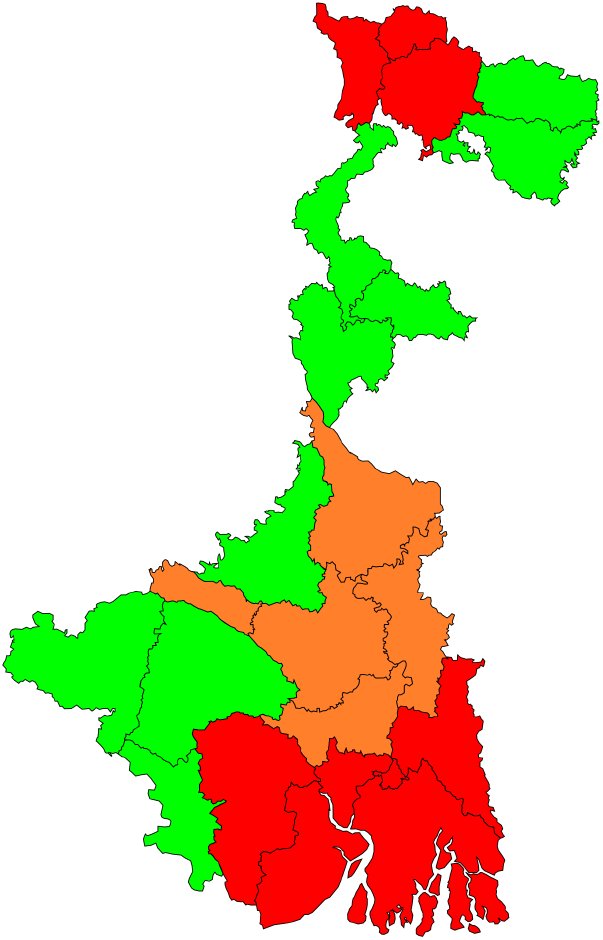
Red,Orange and Green zone of Kolkata with 8 connected districts i.e. East West Bengal[51]. Our selected all districts Medinipur, West Medinipur, Howrah, Hoo-are belonging in red or orange zone by govt. gly, Nadia, Burdwan, North 24 Parganas and notification. South 24 Parganas.

**Figure 3:**
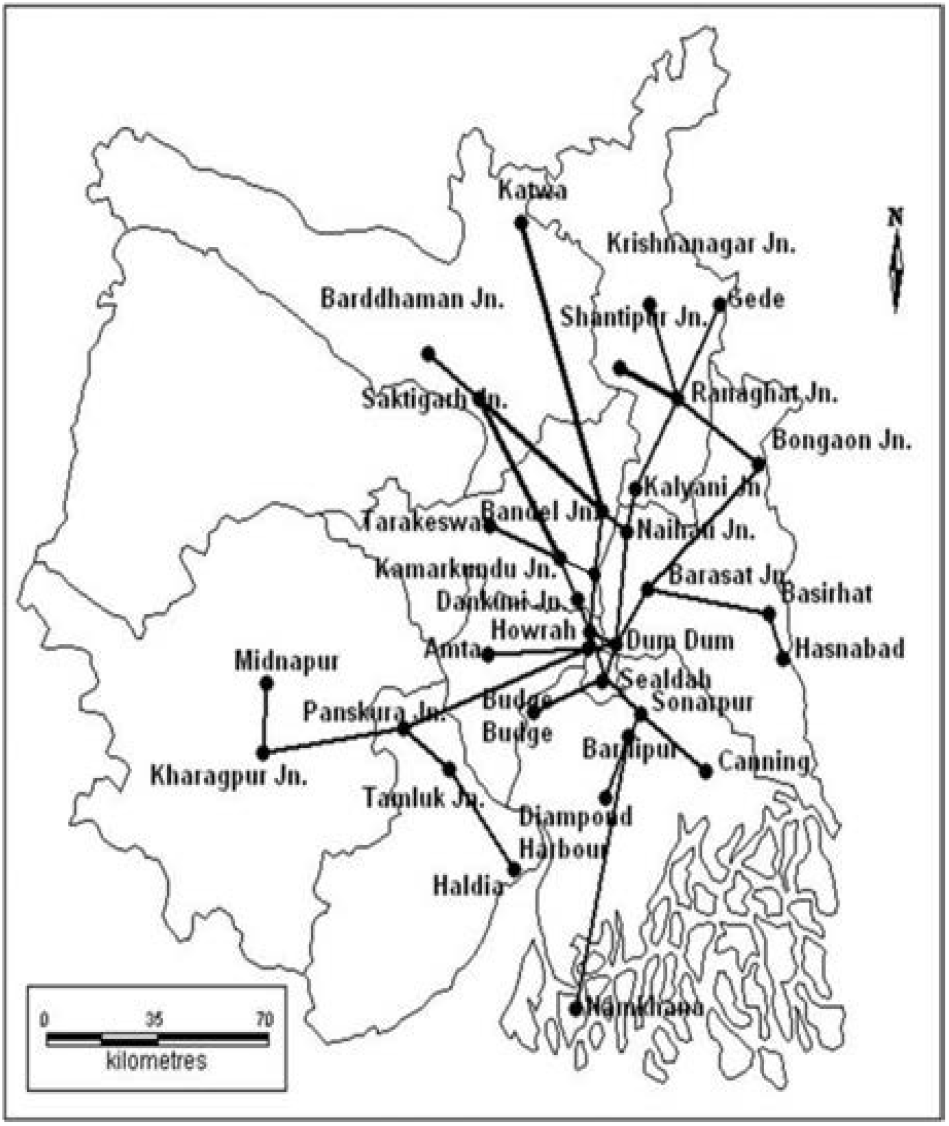
Sub-urban railway map surrounding Kolkata of West Bengal[53]

### 3.2. The selection of regions

West Bengal is a huge and complex state of India: about 10 crore people live here in different conditions. As mentioned earlier, in the state West Bengal, Kolkata is the capital of the province. Kolkata is the financial center for both the

West Bengal and eastern India. The most important industrial belt of the country is a corridor extending north and south of Kolkata along the banks of the Hooghly River [52]. That’s why most of the people from other districts go in want of work or for other purposes to Kolkata on daily base. Thus, we choose Kolkata district as the centre of the network. In order to bridge the existing gap between rural and urban areas, Kolkata has adopted a multimodal transport system where railways play an significant part in raising the regional connectivity between Kolkata and the suburban areas. The districts around Kolkata are connected to the city by an improved suburban rail network (3). The suburban railway areas of Kolkata covers nine districts of South Bengal, for example Burdwan, Nadia, Hooghly, Howrah, East and West Midnapore, North and South 24 Parganas including Kolkata district itself [53]. Therefore the another 8 districts of the network are: East Medinipore(*n*_1_), West Midnapore(*n*_2_), North 24 Parganas(*n*_3_), Howrah(*n*_4_), South 24 Parganas(*n*_5_), Hoogly(*n*_6_), Nadia(*n*_7_) and Burdwan(*n*_8_).

### 3.3. Construction of Flux matrix

Initially for defining the network, we build a flux matrix based on selected districts. Among 9 districts, there are some districts, those are not such a significant spot to be considered as employment center. Therefore, the edges are not bidirectional and for this case the network is asymmetric. Here, we adopt that each node contains a conservation of the residential population, so the essence of the constituents of each row of the matrix is 1. Such a matrix usually describes the societal construction of regions *C, n*_1_,*n*_2_,*n*_3_,*n*_4_,*n*_5_,*n*_6_,*n*_7_,*n*_8_. Here, for constructing the flux matrix some logical assumptions are followed. At the Beginning, let’s presume that very few people living in the central district go to another district for work and we neglect that part. People who live in *n*_1_,*n*_2_,*n*_3_,*n*_4_,*n*_5_,*n*_6_,*n*_7_,*n*_8_ visit the central *C* for doing work. We do not recognize the precise figure of masses who move from one district to another district (besided central city) for work. It is observed as per census report [54] that overall migration percentage is around 15% due to work and employment throughout the country. With this, to fix the values of the parameters to build the flux matrix, we assume the percentage of each vertex that works within the district is 98% and that leaves home for work in others is 2%. Since, inter district migration is not much observed for Kolkata city as per census record, we include intra district employment by fixing *ψ*_11_ =1 and rest of the variables are taken as zero. For more precise study we also employ the data set of Suburban train services of south eastern railway (connecting south, east and north of West Bengal) that includes total number of trains daily piled,their frequencies, number of coaches per train, capacity of each coach and the total number of passengers carried by these trains and finally develop the concerned flux matrix as follows.

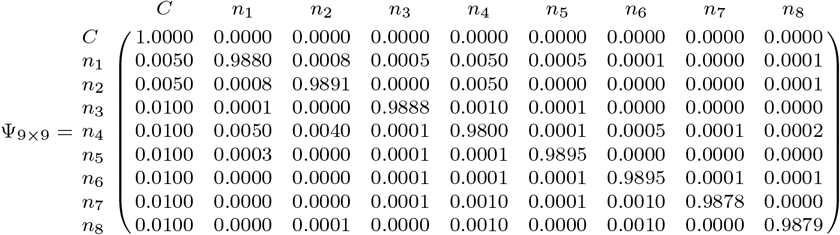

### 3.4. Data source

There are practical difficulties in resolving the actual number of infections, since most cases of covid-19 are non-communicable or the symptoms are influenza-like, for example. Unfortunately, the data obtained only measures the number of people who have visited the hospital for treatment. Thus, we can say that the measurements are significantly below the estimates of the total infected population.

We gather epidemiological data from the following publicly available data sources: covid19india.org: Coronavirus Outbreak in India (https://www.covid19india.org/state/WB) and the Ministry of Health (http://https://www.mohfw.gov.in/).

Here we estimate the transmission co-efficient, recovery rate and disease related mortality based on data from the data sources: covid19india.org: Coronavirus Outbreak in India (https://www.covid19india.org/state/WB) on 25th May,2020.

### 3.5. Estimating epid

In epidemiology, it is a very basic problem to estimate how many new infections occurred during the onset of an epidemic by a single infected person. This number is called the basic reproduction number and denoted by 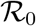. With a sufficient population an epidemic would spread if and only if 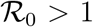. This marginal property has a biological inspiration, and the mathematical theory of 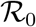 is well described in [48, 21, 47, 42]. As the total number of cases in West Bengal is relatively low, the state has seen a rise in 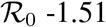 in late April, 1.14 in early May, 1.34 till May 10 and 1.22 in May 15 to May 25 (Source: Data shared by Sitabhra Sinha, Institute of Mathematical Sciences, Chennai, via IndiaSpend)[55]. Now with the mean 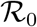 as 1.30, mean viremic period 37 days and normal death rate 0.00002 of West Bengal, we have estimated the recovery rate(*τ*), disease related death(*γ*) and transmission rate(*η*) on 25th May,2020. Also this model represents virus transmission by a set of nonlinear differential equations (ODEs) that add a transmission rate for the dynamics of individuals in compartments. These transmission rate models are used to define a basic reproductive rate 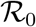 that represents the number of infectious populations that may be infected by an infectious individual. Here for the fixed 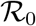 and district wise different recovery and death rate, we have obtained the district wise transmission rate (using the Theorem-1), given in Table-2.

**Table 1:** District wise total and initial susceptible, infected and recovered population

| District | Total Population | Susceptible ($S_i$) | Infected ($I_i$) | Recovered ($R_i$) |
| --- | --- | --- | --- | --- |
| Kolkata | 4486679 | 4484986 | 1693 | 683 |
| Midnapore(E) | 5094238 | 5094176 | 62 | 33 |
| Midnapore(W) | 5943300 | 5943286 | 26 | 19 |
| North 24. Pgs. | 10082852 | 10082372 | 480 | 179 |
| Howrah | 4850029 | 4849217 | 812 | 256 |
| South 24. Pgs. | 8153176 | 8153048 | 128 | 49 |
| Hooghly | 5520389 | 5520173 | 216 | 117 |
| Nodia | 5168488 | 5168416 | 27 | 9 |
| Burdwan(E+W) | 7717563 | 7717504 | 59 | 16 |

**Table 2:** District wise recovery rate, death rate and transmission rate on 25th May

| District | Infected Population(I) | Recovered Population ($R$) | Dead Population ($D$) | Recovery rate ($\tau = R/I$) | Death rate ($\gamma = D/I$) | Transmission rate ($\eta_j$) |
| --- | --- | --- | --- | --- | --- | --- |
| Kolkata(C) | 1693 | 683 | 184 | 0.40 | 0.108 | 0.695 |
| East Midnapore($n_1$) | 62 | 33 | 1 | 0.52 | 0.015 | 0.73 |
| West Midnapore($n_2$) | 28 | 19 | 0 | 0.68 | 0 | 0.919 |
| North 24. Pgs.($n_3$) | 482 | 179 | 38 | 0.37 | 0.078 | 0.617 |
| Howrah($n_4$) | 812 | 256 | 37 | 0.32 | 0.046 | 0.511 |
| South 24. Pgs.($n_5$) | 128 | 49 | 5 | 0.38 | 0.01 | 0.58 |
| Hooghly ($n_6$) | 218 | 117 | 7 | 0.54 | 0.032 | 0.779 |
| Nadia ($n_7$) | 27 | 9 | 0 | 0.33 | 0 | 0.464 |
| Burdwan(E+W) ($n_8$) | 59 | 16 | 2 | 0.27 | 0.033 | 0.429 |

### 3.6. Result analysis

In this network model by exploiting the above data we perform a comparative study in each of these nine cities (districts) of West Bengal. We explore the disease dynamics during and after lock down (i.e. after unlocking) when people start moving from one district to another and form a network for the spread of the SARS-2 or COVID-19. Figures 4 to 21 present the time series plot for susceptible, infectious and recovered population during and after partial lock down restrictions. Figures 22 and 23 compares the time taken by the infected populations of each district to reach to its peak level during partial lock-down and after unlocking respectively. The numerical values are summarized in the Table-3.

**Figure 4:**
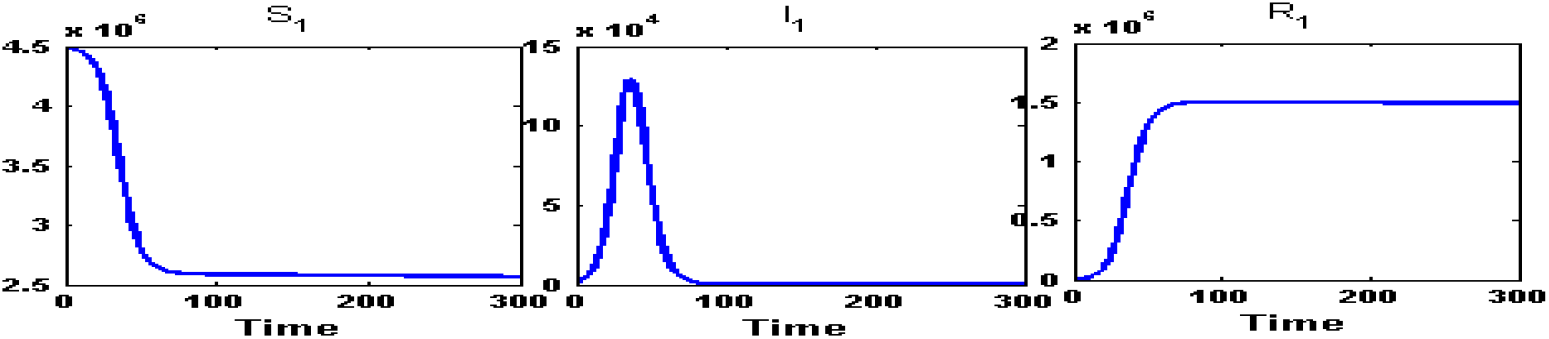
Susceptible (*S*_1_) class, Infected (*I*_1_) class and Recovered (*R*_1_) class people of Kolkata(C) for intra-district network. From an initial infection of an amount 1693 person, during partial lock-down within 37 days the number of infection would have reached at its peak value 1,29,200 and would go below the starting amount at around 79 days.

**Figure 5:**
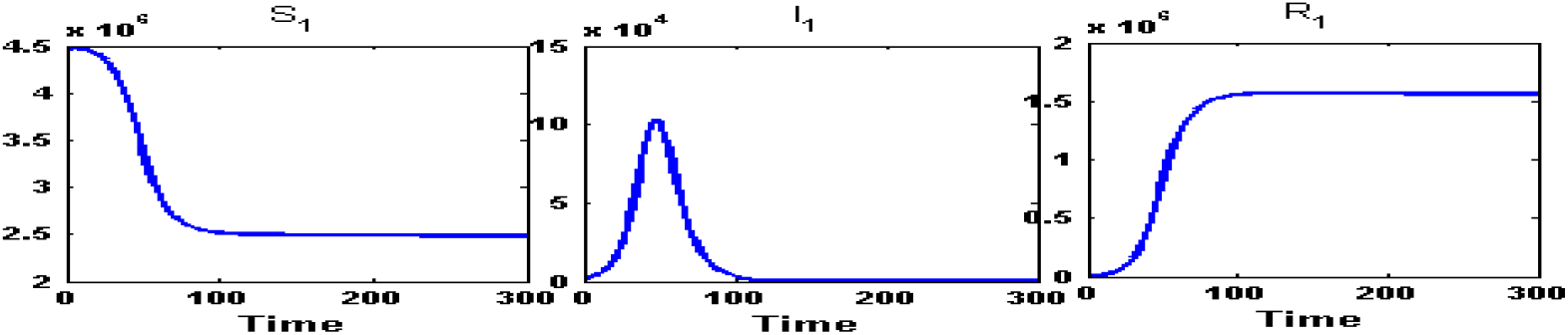
Susceptible (*S*_1_) class, Infected (*I*_1_) class and Recovered (*R*_1_) class people of Kolkata(C) for fully connected network. After the formation of network between these closely related district, the infected population will reach to it’s peak value at an amount 1,03,000 person in 47 days and will go down the initial level of infection in 110 days.

**Figure 6:**
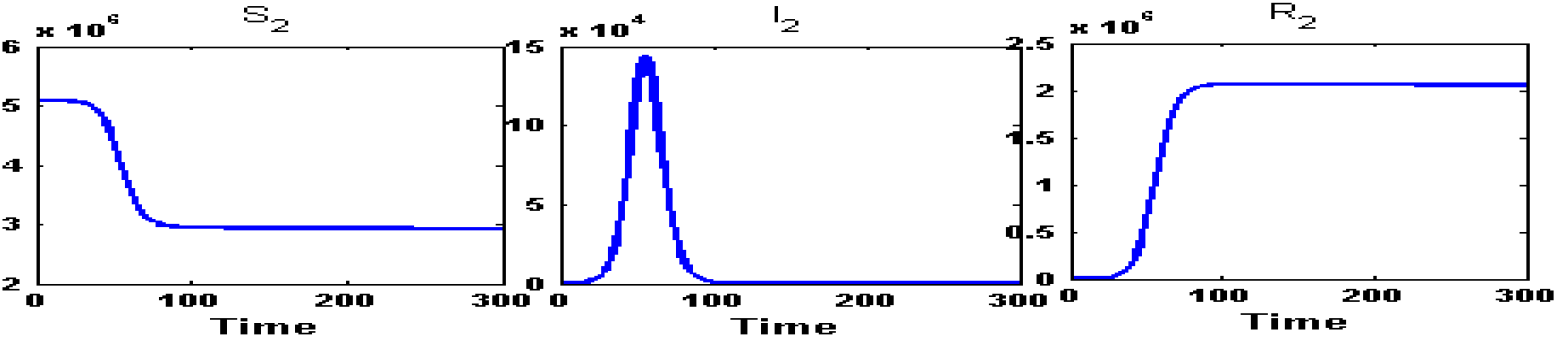
Susceptible (*S*_2_) class, Infected (*I*_2_) class and Recovered (*R*_2_) class people of East Midnapore(*n*_1_) for intra-district network. During the partial lock-down, the infected population will reach at its peak of an amount of 1,43,800 infected person in 56 days and will go below the initial infected in 120 days.

**Figure 7:**
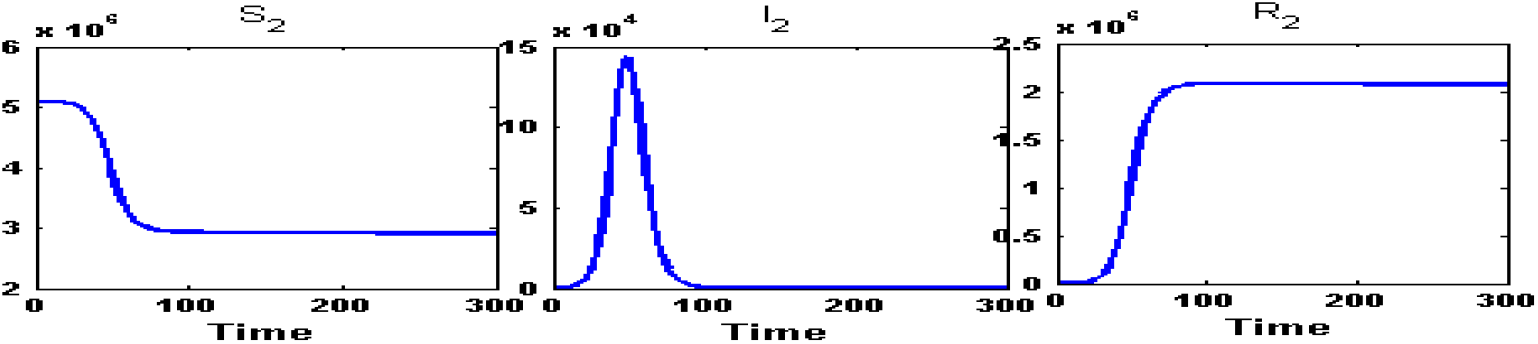
Susceptible (*S*_2_) class, Infected (*I*_2_) class and Recovered (*R*_2_) class people of East Midnapore(*n*_1_) for fully connected network. After the unlocking, within 49 days, the infected population will reach to its peak value at an amount 1,43,700 persons and will reduce down the initial amount of infection in 125 days.

**Figure 8:**
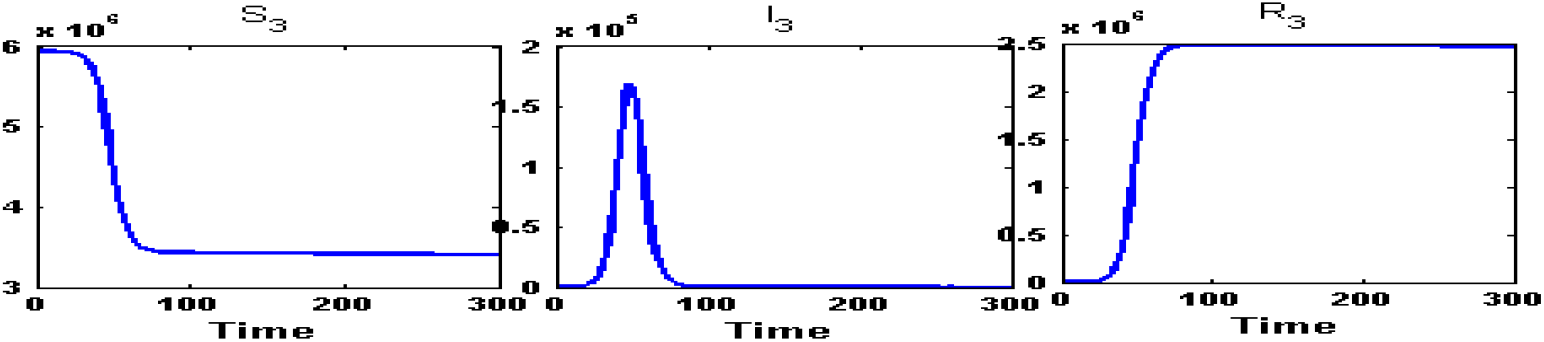
Susceptible (*S*_3_) class, Infected (*I*_3_) class and Recovered (*R*_3_) class people of West Midnapore(*n*_2_) for intra-district network. During the partial lock-down, the infected population will reach at its peak of an amount of 1,68,900 infected person in 48 days and will go below the initial infected in 127 days.

**Figure 9:**
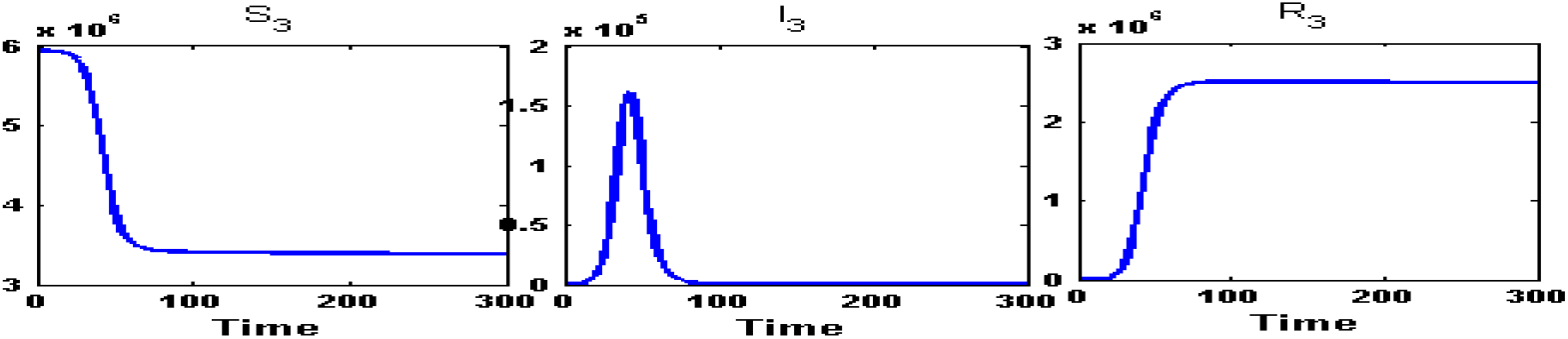
Susceptible (*S*_3_) class, Infected (*I*_3_) class and Recovered (*R*_3_) class people of West Midnapore(*n*_2_) for fully connected network. After the unlocking, within 43 days, the infected population will reach to its peak value at an amount 1,61,300 persons and down the initial amount of infection in 128 days.

**Figure 10:**
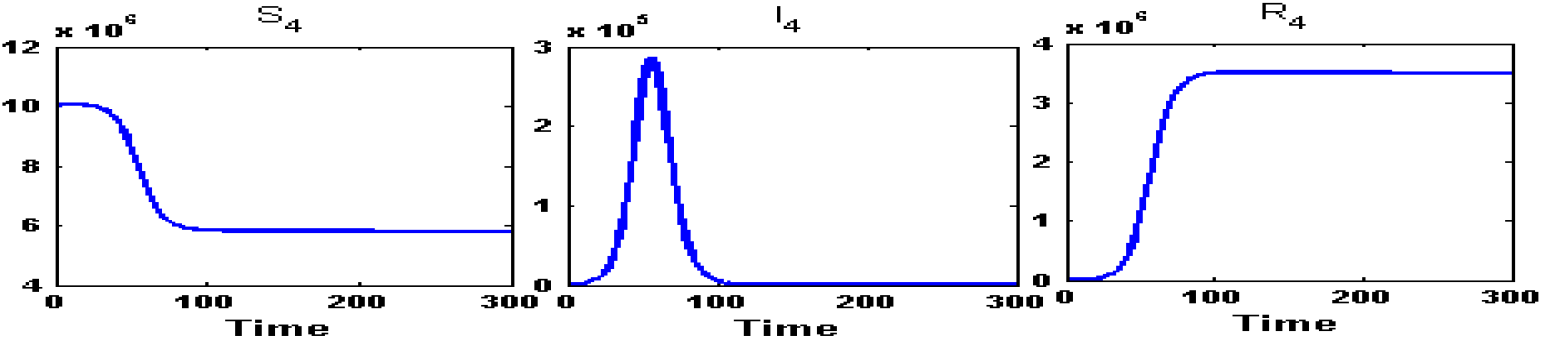
Susceptible (*S*_4_) class, Infected (*I*_4_) class and Recovered (*R*_4_) class people of North 24 Parganas(*n*_3_) for intra-district network. During the partial lock-down, the infected population will reach at its peak of an amount of 2,86,100 infected person in 56 days and will go below the initial infected in 122 days.

**Figure 11:**
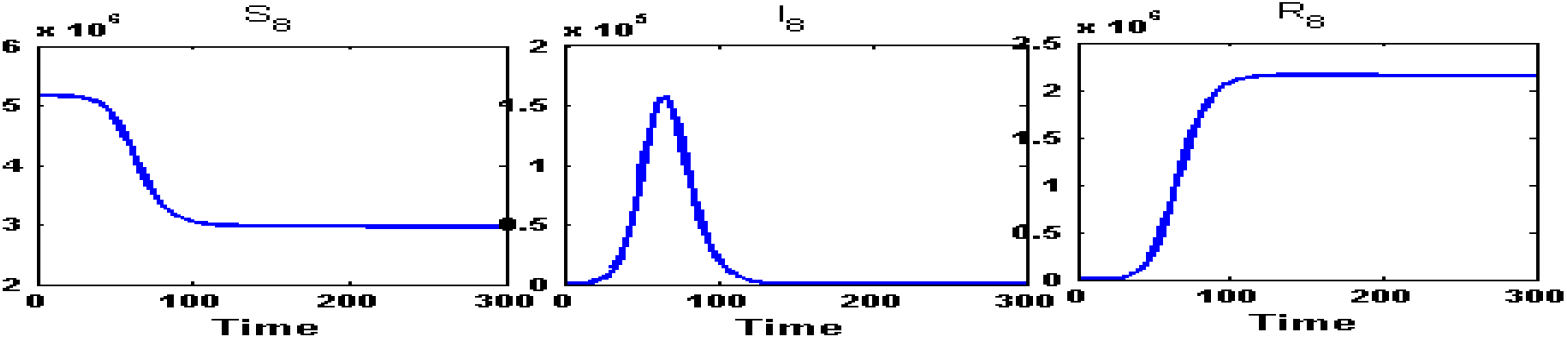
Susceptible (*S*_4_) class, Infected (*I*_4_) class and Recovered (*R*_4_) class people of North 24 Parganas(*n*_3_) for fully connected network. After the unlocking, within 51 days, the infected population will reach to its peak value at an amount 2,90,400 persons and down the initial amount of infection in 119 days.

**Figure 12:**
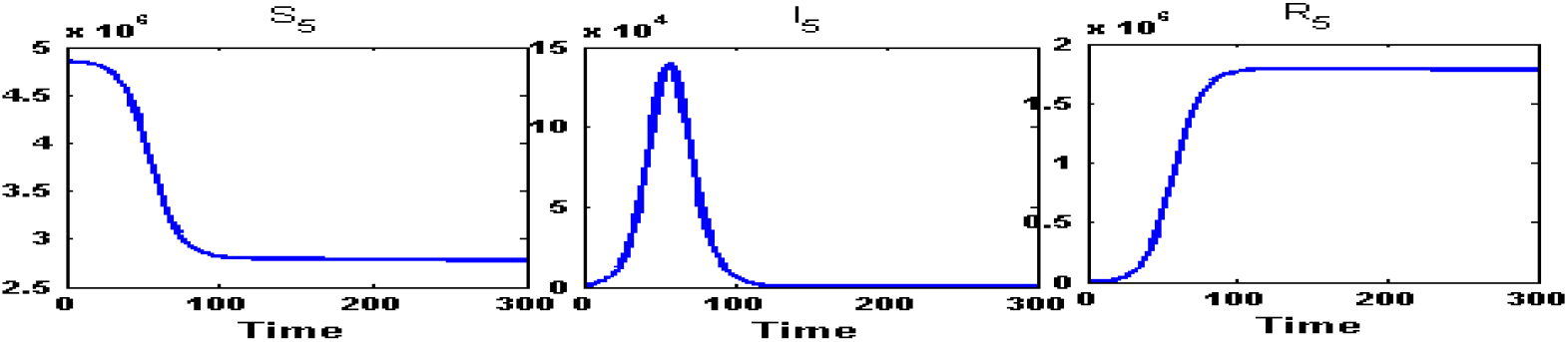
Susceptible (*S*_5_) class, Infected (*I*_5_) class and Recovered (*R*_5_) class people of Howrah(*n*_4_) for intra-district network. During the partial lock-down, the infected population will reach at its peak of an amount of 1,39,200 infected person in 56 days and will go below the initial infected in 122 days.

**Figure 13:**
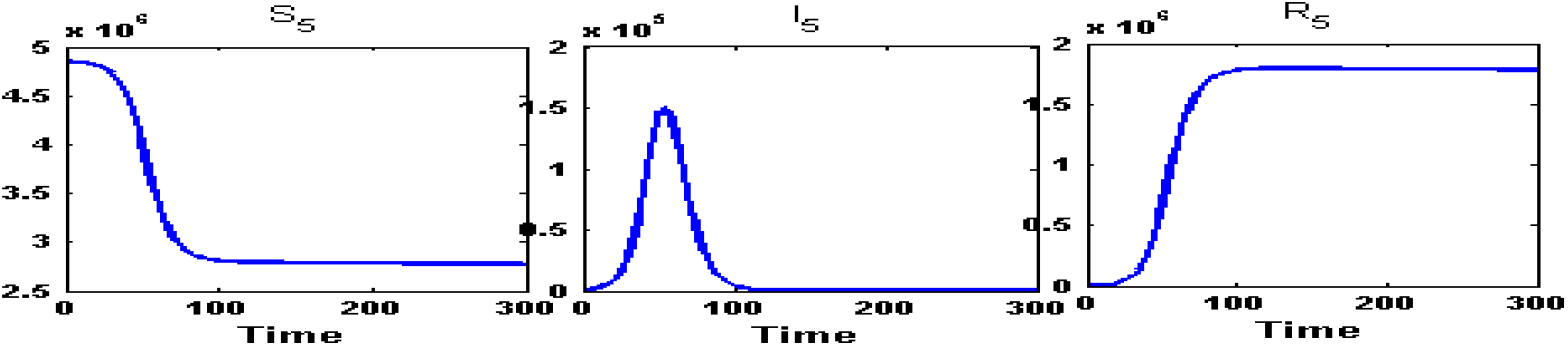
Susceptible (*S*_5_) class, Infected (*I*_5_) class and Recovered (*R*_5_) class people of Howrah(*n*_4_) for fully connected network. After the unlocking, within 53 days, the infected population will reach to its peak value at an amount 1,50,100 persons and down the initial amount of infection in 119 days.

**Figure 14:**
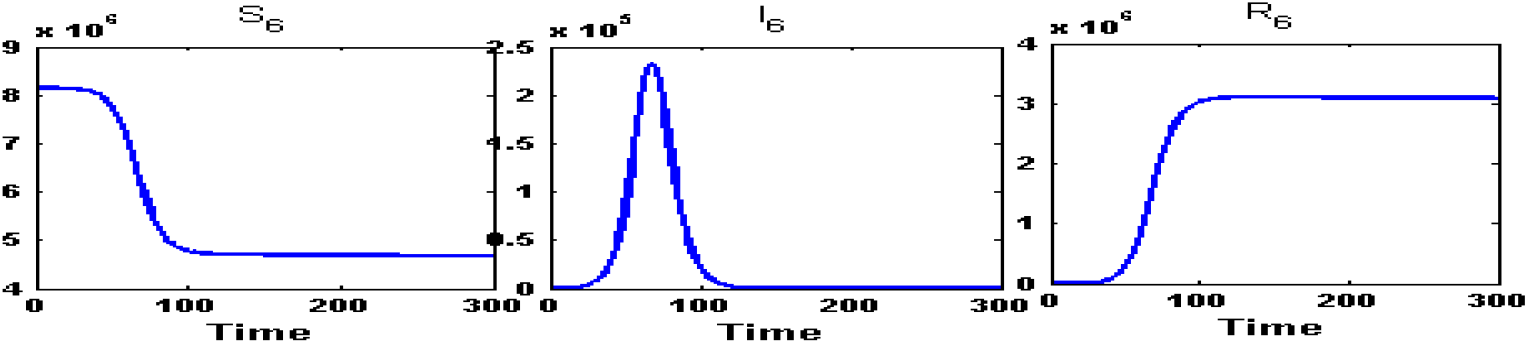
Susceptible (*S*_6_) class, Infected (*I*_6_) class and Recovered (*R*_6_) class people of South 24 Parganas(*n*_5_) for intra-district network. During the partial lock-down, the infected population will reach at its peak of an amount of 2,32,400 infected person in 68 days and will go below the initial infected in 147 days.

**Figure 15:**
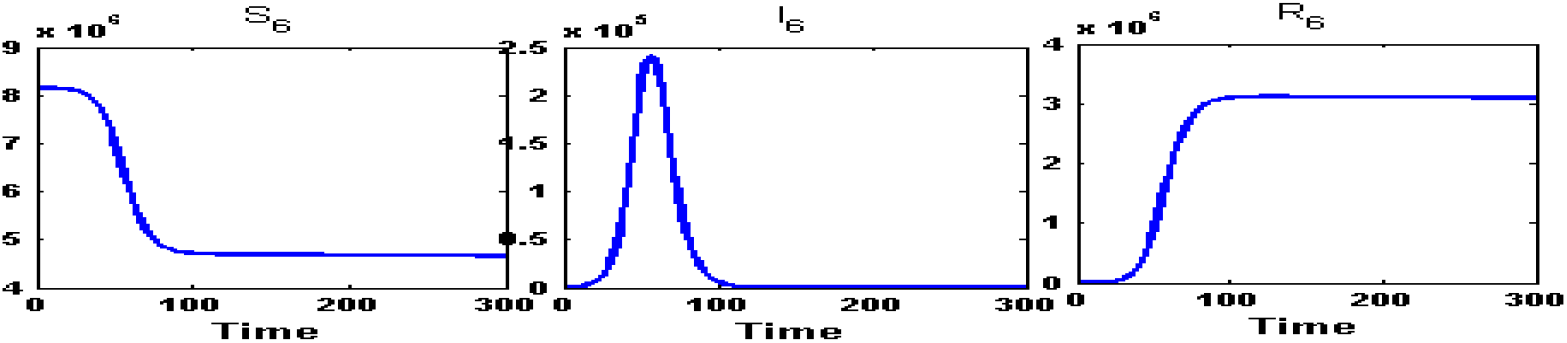
Susceptible (*S*_6_) class, Infected (*I*_6_) class and Recovered (*R*_6_) class people of South 24 Parganas(*n*_5_) for fully connected network. After the unlocking, within 58 days, the infected population will reach to its peak value at an amount 2,39,100 persons and down the initial amount of infection in 137 days.

**Figure 16:**
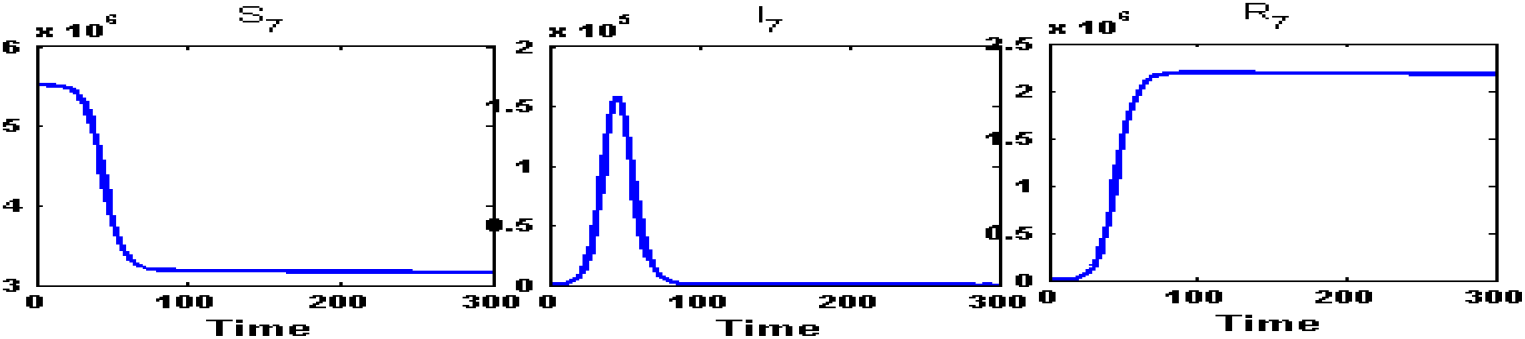
Susceptible (*S*_7_) class, Infected (*I*_7_) class and Recovered (*R*_7_) class people of Hooghly(*n*_6_) fot intra-district network. During the partial lock-down, the infected population will reach at its peak of an amount of 1,57,900 infected person in 45 days and will go below the initial infected in 98 days.

**Figure 17:**
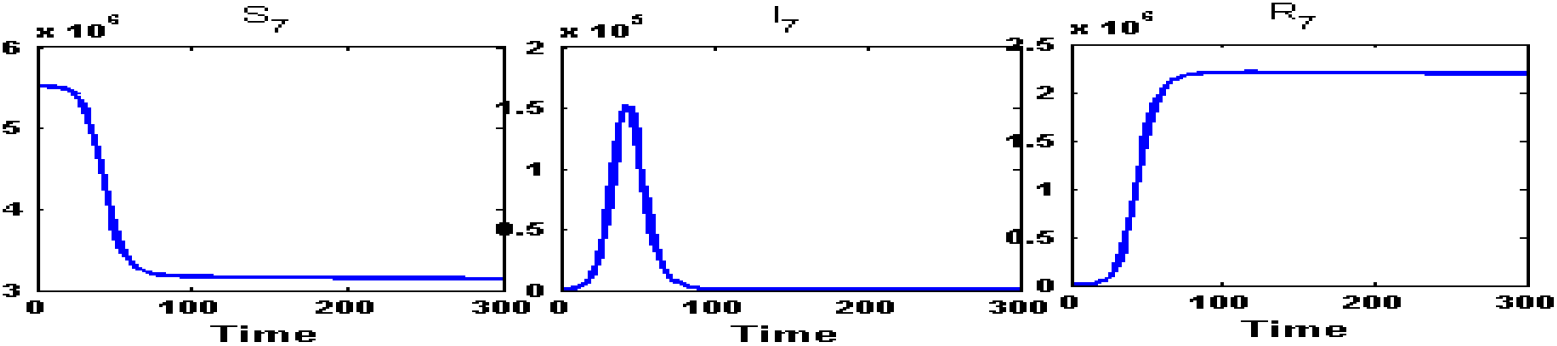
Susceptible (*S*_7_) class, Infected (*I*_7_) class and Recovered (*R*_7_) class people of Hooghly(*n*_6_) fot fully connected network. After the unlocking, within 43 days, the infected population will reach to its peak value at an amount 1,52,500 persons and down the initial amount of infection in 116 days.

**Figure 18:**
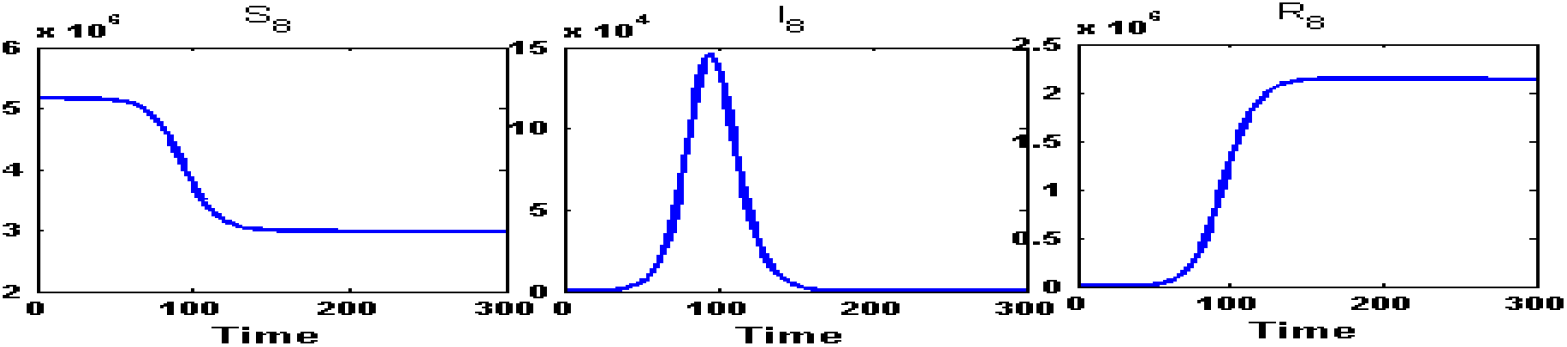
Susceptible (*S*_8_) class, Infected (*I*_8_) class and Recovered (*R*_8_) class people of Nadia(*n*_7_) for intra-district network. During the partial lock-down, the infected population will reach at its peak of an amount of 1,45,400 infected person in 96 days and will go below the initial infected in 207 days.

**Figure 19:**
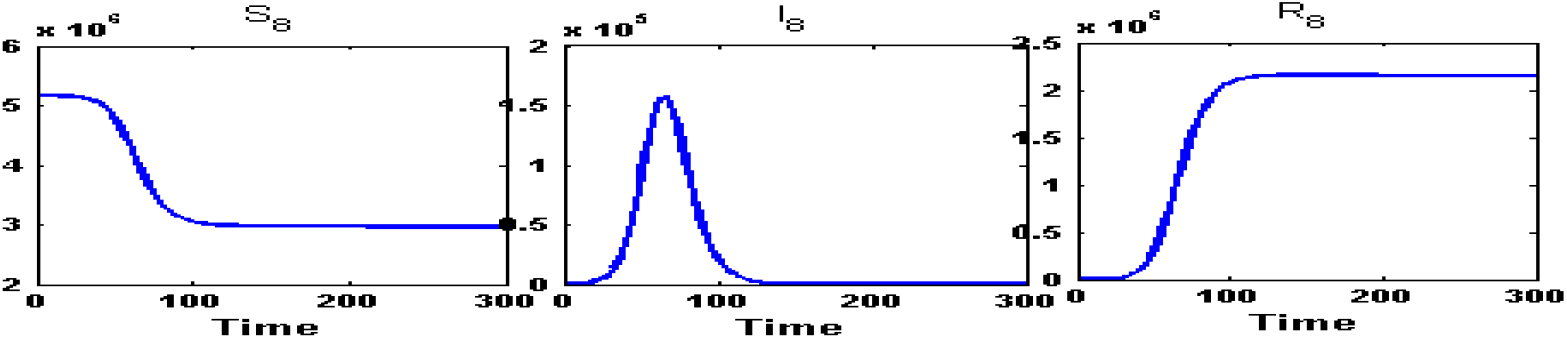
Susceptible (*S*_8_) class, Infected (*I*_8_) class and Recovered (*R*_8_) class people of Nadia(*n*_7_) for fully connected network. After the unlocking, within 66 days, the infected population will reach to its peak value at an amount 1,57,100 persons and down the initial amount of infection in 175 days.

**Figure 20:**
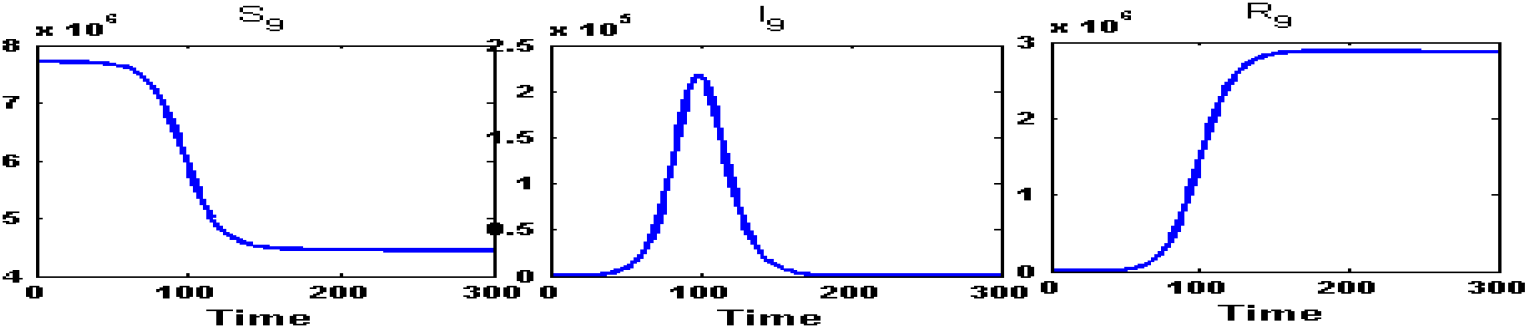
Susceptible (*S*_9_) class, Infected (*I*_9_) class and Recovered (*R*_9_) class people of Burdwan(*n*_8_) for intra-connected network. During the partial lock-down, the infected population will reach at its peak of an amount of 2,17,400 infected person in 100 days and will go below the initial infected in 215 days

**Figure 21:**
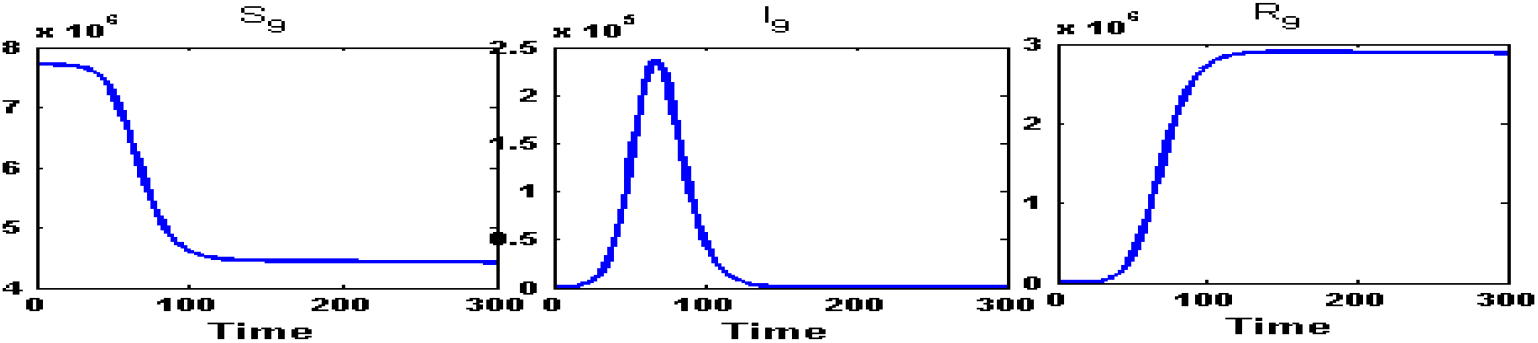
Susceptible (*S*_9_) class, Infected (*I*_9_) class and Recovered (*R*_9_) class people of Burdwan(*n*_8_) for fully connected network. After the unlocking, within 69 days, the infected population will reach to its peak value at an amount 2,36,600 persons and down the initial amount of infection in 182 days.

**Figure 22:**
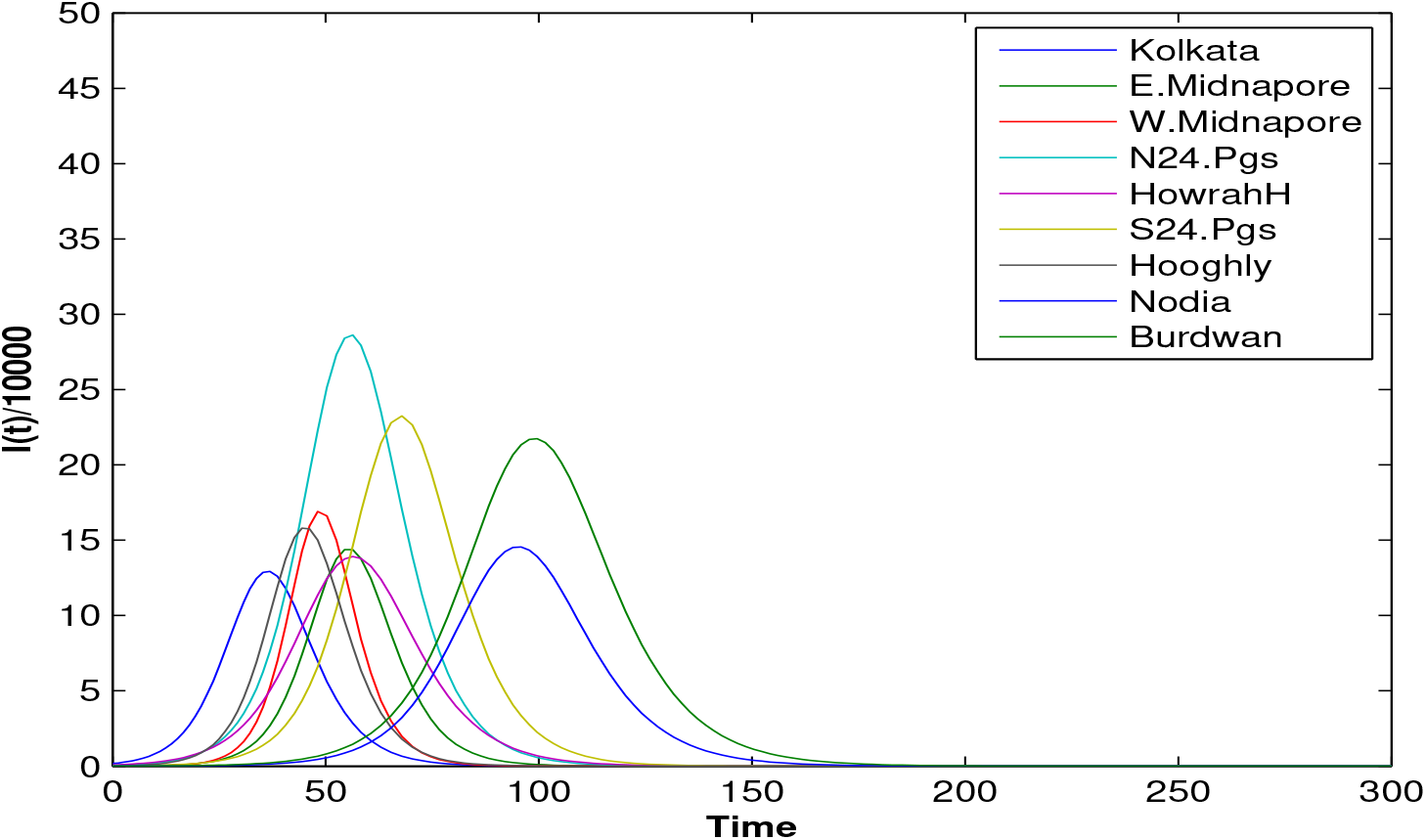
Infected population when the transportation open within their own network (Time Vs *I_i_/*10000). Infected population to reach at its peak for each of the nine districts will range between 37 to 100 days

**Figure 23:**
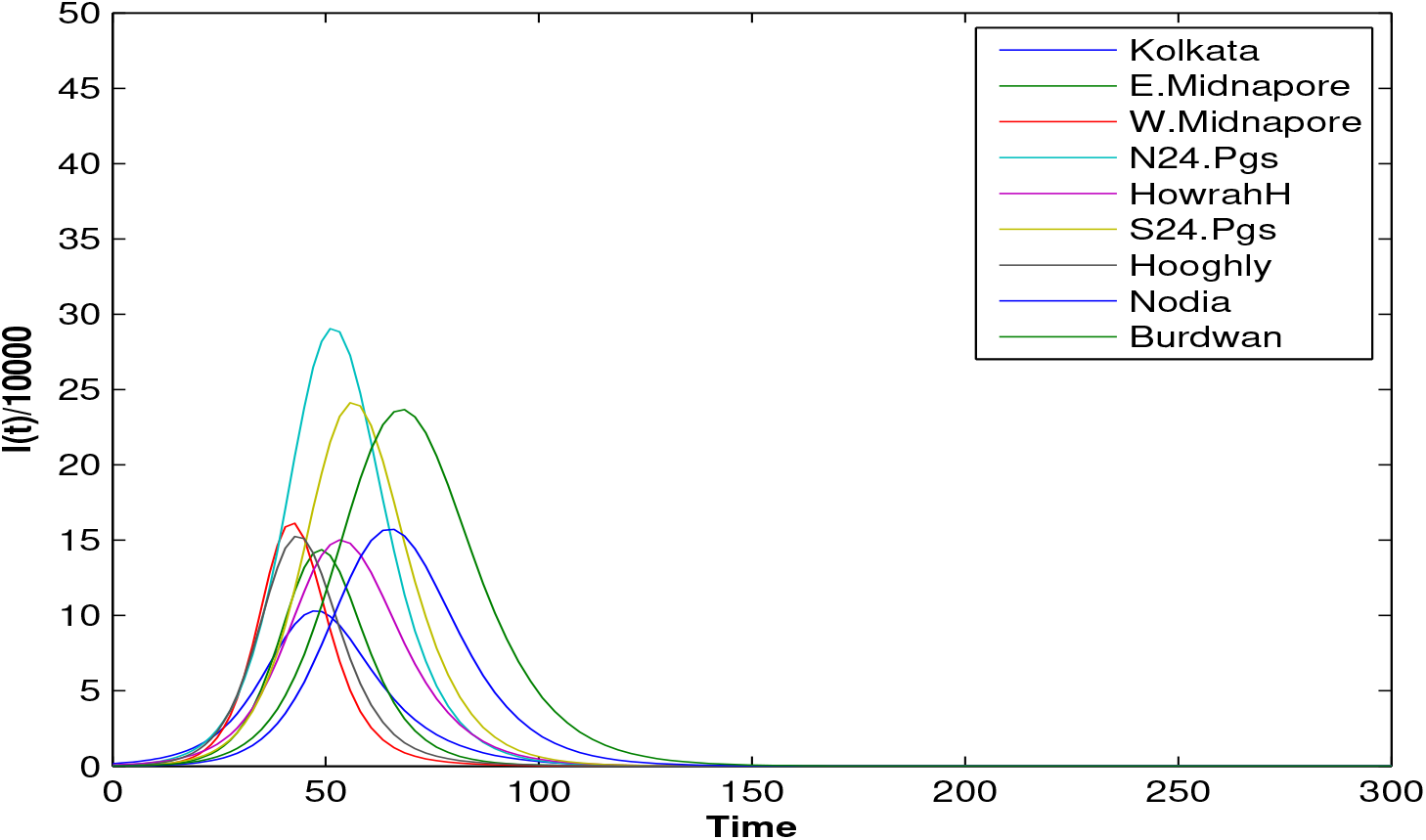
Infected population when the transportation open within the full network (Time Vs *I_i_/*10000). Infected population to reach at its peak for each of the nine districts will range between 43 to 69 days

**Table 3:** Infected population summary during partial lock-down and after unlock

| District | Initial Infection | During partial lock-down |  |  |  | After unlock |  |  |  |
| --- | --- | --- | --- | --- | --- | --- | --- | --- | --- |
|  |  | Peak | Days | Below Initial | Days | Peak | Days | Below Initial | Days |
| Kolkata | 1693 | 129200 | 37 | 1596 | 79 | 103000 | 47 | 1348 | 110 |
| Midnapore(E) | 62 | 143800 | 56 | 58 | 120 | 143700 | 49 | 55 | 125 |
| Midnapore(W) | 26 | 168900 | 48 | 20 | 127 | 161300 | 43 | 26 | 128 |
| North 24 Pgs. | 480 | 286100 | 56 | 398 | 122 | 290400 | 51 | 467 | 119 |
| Howrah | 812 | 139200 | 56 | 768 | 122 | 150100 | 53 | 763 | 119 |
| South 24 Pgs. | 128 | 232400 | 68 | 124 | 147 | 239100 | 58 | 113 | 137 |
| Hooghly | 216 | 157900 | 45 | 215 | 98 | 152500 | 43 | 187 | 116 |
| Nadia | 27 | 145400 | 96 | 25 | 207 | 157100 | 66 | 23 | 175 |
| Burdwan(E+W) | 59 | 217400 | 100 | 56 | 215 | 236600 | 69 | 50 | 182 |

It can be seen from all the figures that, most of the susceptible class individuals have been entered into infected class after some days. It also shows that most people are recovering after reaching the saturation level of the infected class.

## 4. Discussion

Here we have used SIR-network Predictive model in two circumstances, during and after partial lock-down, to generate dynamical predictions and help inform the debate as to easing lock-down measures. Just like any other model, our model also suffers from deviations in accuracy in predicting the spread and peak of the outbreak. These deviations can be attributed to factors like alternative release situations and such other border conditions. None the less we have provided the model scenarios that can serve as guidelines in the fight against the COVID-19 pandemic. Furthermore, in unlocking the populations newer spread patterns of SARS-COV-2 will be observed. So focus in this work is on dynamic modeling of this pandemic so that we can fit in most of future lock-down scenarios in mathematically cautious fashion and as such we have formulated our model in such a way that at the same time as having isolated parameters it is complex enough to capture a wide range of options. Each other parameters relate either to the level of infection or to the types and variety of situations that prevailed before a lock-down is affected. In order to deal with the inadequacies of the knowledge of COVID-19 we have brought some changes in the basic parameters so that an optimal strategy can effectively be handled. Graphical analysis shows that amongst the nine districts included in this study, the dynamics of COVID-19 infection patterns cannot be generalized. In all districts more or less the peak is reached earlier than the lock-down situation. Except in the capital city Kolkata where the peak is reached much lesser time. This can be explained in the fact that migration happens from the districts to Kolkata and not vice-versa raising the number of incumbent population in the city and hence raising the number of infections. In the other districts also the results are varied. In West Midnapore and Hoogly for example the peak is reached much earlier in a partial lock-down situation that is an unlocked situation due to the high rate of infection. In East Midnapore the peak situation is more or less same in all scenarios because of the high rate of travel whereas in Burdwan and Nadia the peak is reached after a longer time than the other districts because of high rate of recovery and low rate of infection. It is also observed that people naturally tend to migrate from highly infected areas to low infection areas. We can make one conclusion from here. In all the districts where the number of infected people is increasing after the introduction of full network resume, partial network should be initiated on those contaminated zones and full network should be imposed on the rest of the less infected areas.

It is generally observed that after the resumption of transportation the total number of infections has fallen and as such our results indicate that the overall situation can improve if the policy makers allow partial reopening of transport networks in contentment areas while allowing full access to travel in the other less affected areas. We also come to observe that in a densely populated country like India the risk of infection will always be unavoidably high and the initial 90 days lock-down has already proven to be a setback for the economy not just India but the entire world has suffered. Thus, with services and manufacturing breaking down in several places and Global supply chains breaking down also. As an example, the work from home scenario in India has hit hard the transporters all round the country and whatever little is available is actually exorbitantly priced of those commuters who are compelled to travel even in the pandemic. Aspiring students mentally distort because of the postponing of their exams and the prospect for jobs being rendered slightly uncertain not to mention the raising labor downsizing that these youngsters are observing around them. With an effective vaccine being out of which at the moment, it is difficult for the policy makers to find balance between saving, the economy and saving lives from COVID-19 and puts additional pressure on health care and every other human enterprise. In our work, we are trying to tackle this dilemma by considering both the partial lock-down and the unlock models and we have the following assumptions and observations.

- The coefficient of transmission has been fixed in the calculation. However, it is hard to retain it when unlock leads to migration, so that in order to accommodate this scenario, social distancing has to be maintained everywhere.
- In the absence of both a readily available COVID-19 vaccine and adequate centralized medical facilities available for everybody, medicine must be administered locally, including in home isolation, so that treatment may be meted out to all affected, as such potable medical units and instruments must be accrued. This can be arranged during any of the subsequent lock-down periods when the stress of handling large number of patients is low.
- If after gathering enough medical facilities for both centralized and localized treatments the general population is allowed to lead a normal life, the increase number of infections will make it to the peak point of the pandemic in India after which the number of cases is going to start to drop. To be specific our study shows that from the time of unlocking the number of infections will start to drop after 4 months.
- When we started to study the results after the simplification of the lock-down system, we observe that recovery rate is one of the most important parameters in understanding post infection peaks. Similarly, specific studies can also be undertaken with other sensitive parameters like infection rates. As the values are analyzed and understood more and more the trustworthiness of our calculation will go on increasing.

Due to complexity of COVID-19 transmissions, the spread of asymptomatic cases has always been a point of concern. As routine rapid testing is impossible to perform in India due to its population or even if it is planned to do so then also it might take approximately 29 years to complete the process. Most of the tests are being performed here for those who possess basic symptoms and those who have come in contact with them. Thus a number of asymptomatic patients are also being identified but that number is very low. This is because the simulated data in the case of analyzing the situations is quite large as compared to real time scenario. The model would be more realistic if the infected population could be divided into several compartments but in that case again there is a problem of data availability for India. From the discussion it is very clear that the SIR-network model is a perfect set of work to be carried out in case of analyzing SARS CoV 2 in India for statistical evaluation of COVID-19 infected population by taking both symptomatic and asymptomatic cases together.

Finally we have validated the model with data from July 21,2020 and the results are given in Appendix. We have found that it took almost the same time to reach the saturation level of the infected people of the every district of our model.

## 5. Conclusion

In conclusion, we can say that as we have evidence from Fig. 4 to Fig. 21, an extended lock-down does not bid well for the economy at all and the indicators also show that extended lock-down does not lower the risk of infection or reinfection. However, extended lock-down will definitely go towards hampering the economy to a large extent and this effect can be neutralized by a period of unlocking. This period of unlocking will help not only in reviving the economy, but it will also help to reach the saturation point of the pandemic in West Bengal in particular or India in general sooner than in the lock-down scenario. That is to say the indicators in the lock-down scenario when analyzed speak of a saturation point which is more uncertain and lengthier than the saturation point visible under unlocking conditions. Now here we must say that if a vaccine is invented or if we are to use a life saving setup like a ventilator, situations will vary. However, in case of lengthy infections that is infections, which last a long time use of ventilators may not be that advisable because ventilators may in some case aid in the collapsing of the lungs rather than reviving the patient. This happens in case of poor compatibility under ventilated conditions in the lungs. So, since ventilators are a very common tool in fighting the Corona virus infection. It is advisable that some intelligent algorithms to be used while using the ventilator that taking to account factors like FRC of the patient compatibility of the patient’s lungs with respect to the ventilator that has been used and so on. At present we should use stochastic modelling to better understand beyond the set of the second web of infections regarding our sample population. However, our SIR model that we have used can easily be adopted to determine the time frame up to the second wave of transmission in the probability setting for better evaluation.

Our model is broad enough to allow it to be applied in countries outside the India or the province West Bengal, starting only with alternative initial conditions and setting certain parameters that specify the country. This must be supported by an agreed uniform definition of our basic model parameters and a case confirmation definition to ensure that the model validity is consistent with the concerned country. In the present situation stochastic frameworks might be more suitable for modelling the exact time period when the population resumes, so the scope of the time frame up to the second wave of transmission in the probability setting can be better evaluated.

## Data Availability

There are practical difficulties in resolving the actual number of infections,
since most cases of covid-19 are non-communicable or the symptoms are influenzalike,
for example. Unfortunately, the data obtained only measures the number of
people who have visited the hospital for treatment. Thus, we can say that the measurements
are significantly below the estimates of the total infected population.
We gather epidemiological data from the following publicly available data
sources: covid19india.org: Coronavirus Outbreak in India (https://www.covid19india.org/state/WB)
and the Ministry of Health (http://https://www.mohfw.gov.in/).

https://www.mohfw.gov.in

## Compliance with Ethical Standards

### Funding

None

### None Conflict of Interest

The authors declare that they have no conflict of interest.

## Appendix A. Appendix

**Table A.4:**
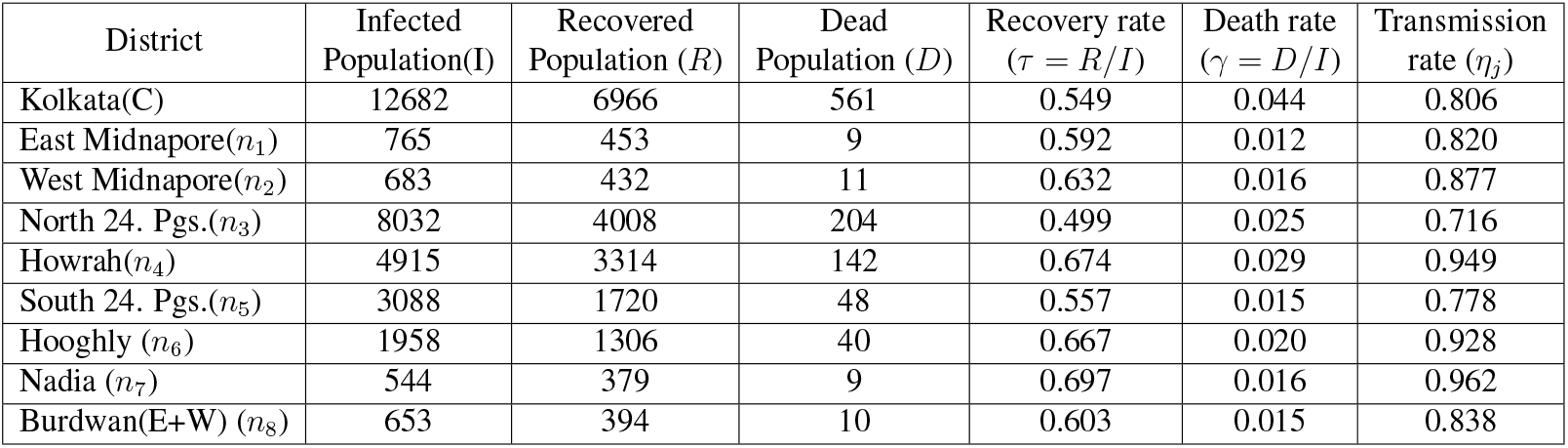
District wise recovery rate, death rate and transmission rate on 18th July,2020

| District | Infected Population(I) | Recovered Population (R) | Dead Population (D) | Recovery rate ($\tau = R/I$) | Death rate ($\gamma = D/I$) | Transmission rate ($\eta_j$) |
| --- | --- | --- | --- | --- | --- | --- |
| Kolkata(C) | 12682 | 6966 | 561 | 0.549 | 0.044 | 0.806 |
| East Midnapore($n_1$) | 765 | 453 | 9 | 0.592 | 0.012 | 0.820 |
| West Midnapore($n_2$) | 683 | 432 | 11 | 0.632 | 0.016 | 0.877 |
| North 24. Pgs.($n_3$) | 8032 | 4008 | 204 | 0.499 | 0.025 | 0.716 |
| Howrah($n_4$) | 4915 | 3314 | 142 | 0.674 | 0.029 | 0.949 |
| South 24. Pgs.($n_5$) | 3088 | 1720 | 48 | 0.557 | 0.015 | 0.778 |
| Hooghly ($n_6$) | 1958 | 1306 | 40 | 0.667 | 0.020 | 0.928 |
| Nadia ($n_7$) | 544 | 379 | 9 | 0.697 | 0.016 | 0.962 |
| Burdwan(E+W) ($n_8$) | 653 | 394 | 10 | 0.603 | 0.015 | 0.838 |

**Figure A.24:**
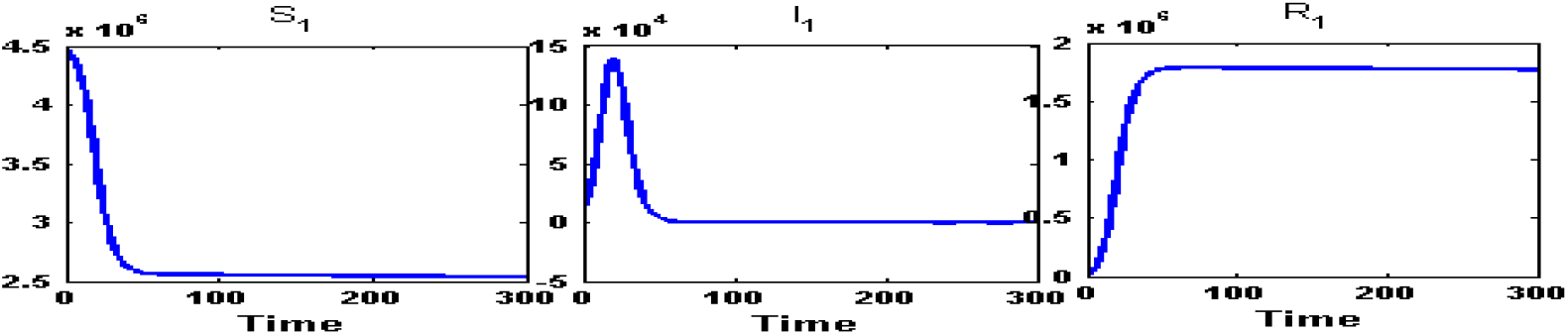
Susceptible (*S*_1_) class, Infected (*I*_1_) class and Recovered (*R*_1_) class people of Kolkata(C) for intra-district network. From an initial infection of an amount 12682 person on 18th July, during partial lock-down within 21 days the number of infection would have reached at its peak value 1,38,700 and would go below the starting amount at around 44 days.

**Figure A.25:**
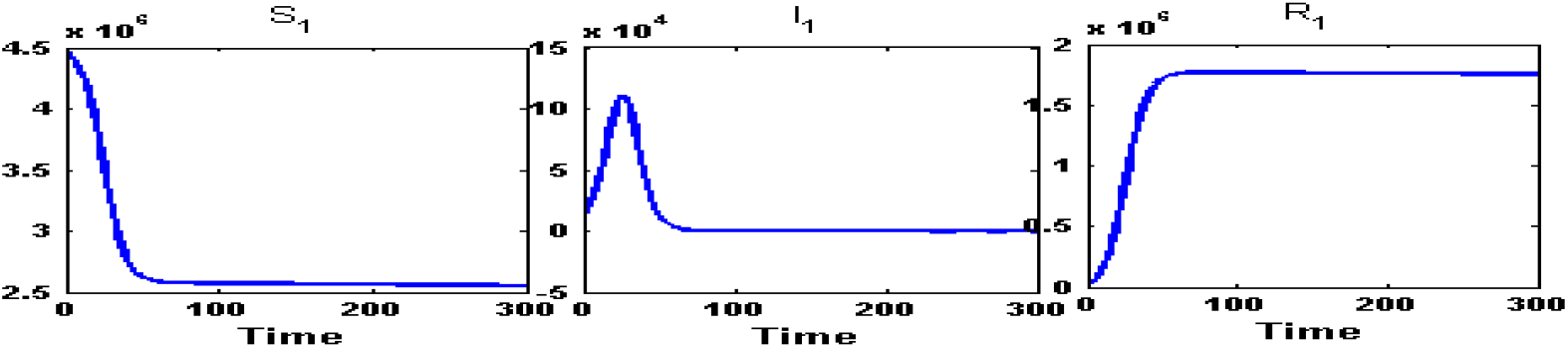
Susceptible (*S*_1_) class, Infected (*I*_1_) class and Recovered (*R*_1_) class people of Kolkata(C) for fully connected network. After the formation of network between these closely related district, the infected population will reach to it’s peak value at an amount 1,10,900 person in 26 days and will go down the initial level of infection in 51 days.

**Figure A.26:**
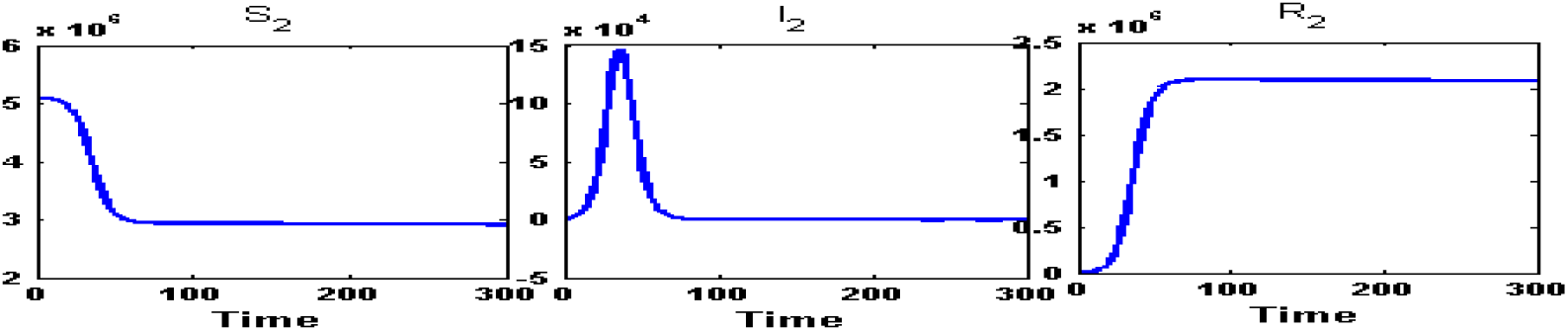
Susceptible (*S*_2_) class, Infected (*I*_2_) class and Recovered (*R*_2_) class people of East Midnapore(*n*_1_) for intra-district network. During the partial lock-down, the infected population will reach at its peak of an amount of 1,46,000 infected person in 37 days from the initial infected population of the amount 765 and will go below the initial infected in 79 days.

**Figure A.27:**
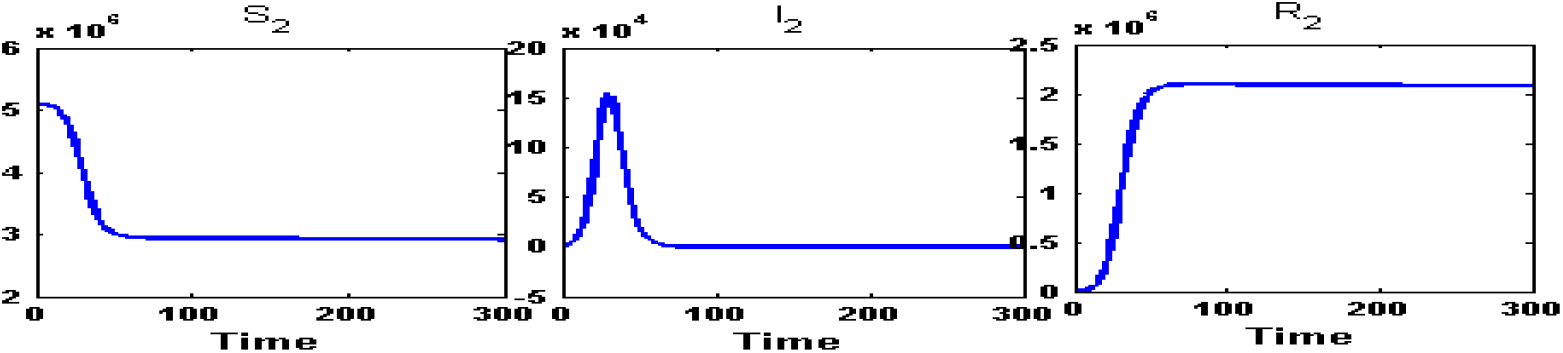
Susceptible (*S*_2_) class, Infected (*I*_2_) class and Recovered (*R*_2_) class people of East Midnapore(*n*_1_) for fully connected network. After the unlocking, within 30 days, the infected population will reach to its peak value at an amount 1,53,800 persons and will reduce down the initial amount of infection in 72 days.

**Figure A.28:**
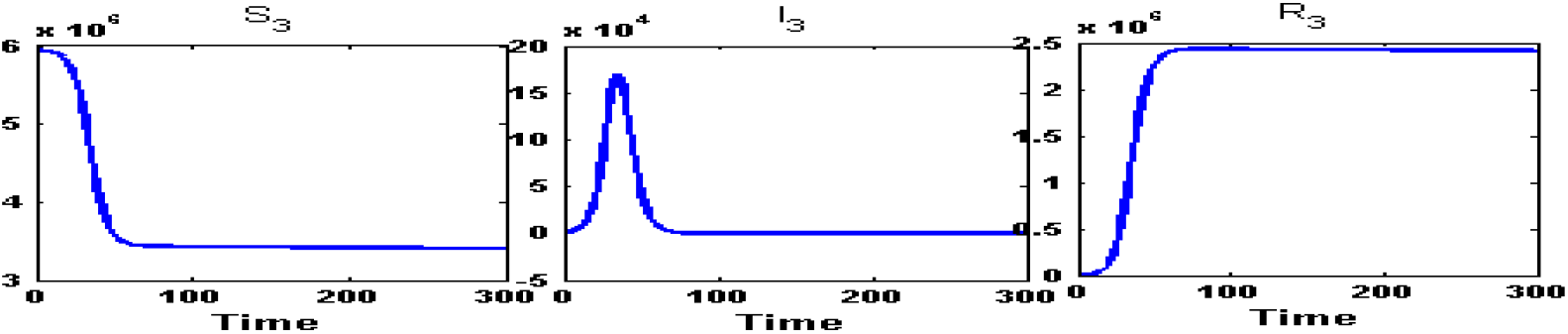
Susceptible (*S*_3_) class, Infected (*I*_3_) class and Recovered (*R*_3_) class people of West Midnapore(*n*_2_) for intra-district network. During the partial lock-down, the infected population will reach at its peak of an amount of 1,70,200 infected person in 35 days from the initial infected population of the amount 683 and will go below the initial infected in 75 days.

**Figure A.29:**
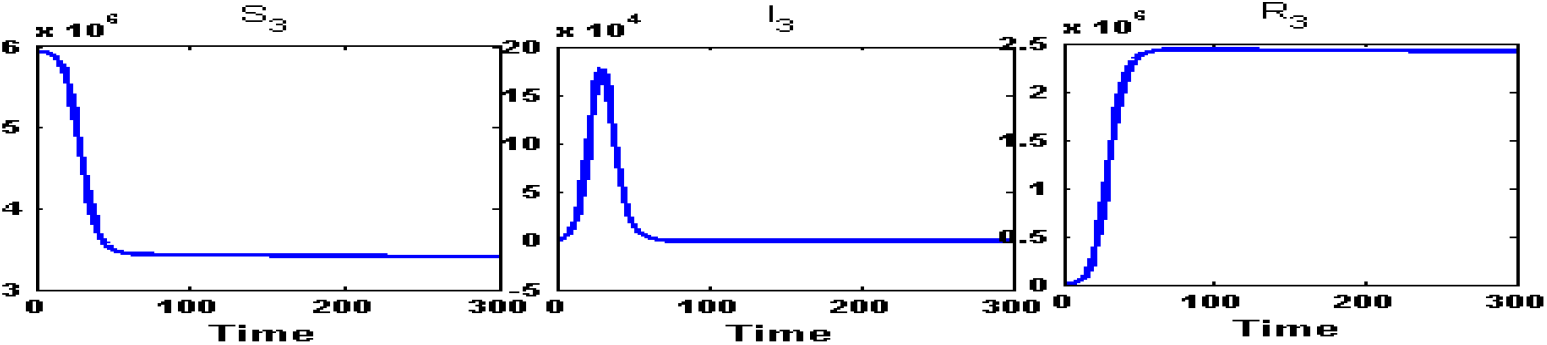
Susceptible (*S*_3_) class, Infected (*I*_3_) class and Recovered (*R*_3_) class people of West Midnapore(*n*_2_) for fully connected network. After the unlocking, within 43 days, the infected population will reach to its peak value at an amount 1,77,300 persons and down the initial amount of infection in 70 days.

**Figure A.30:**
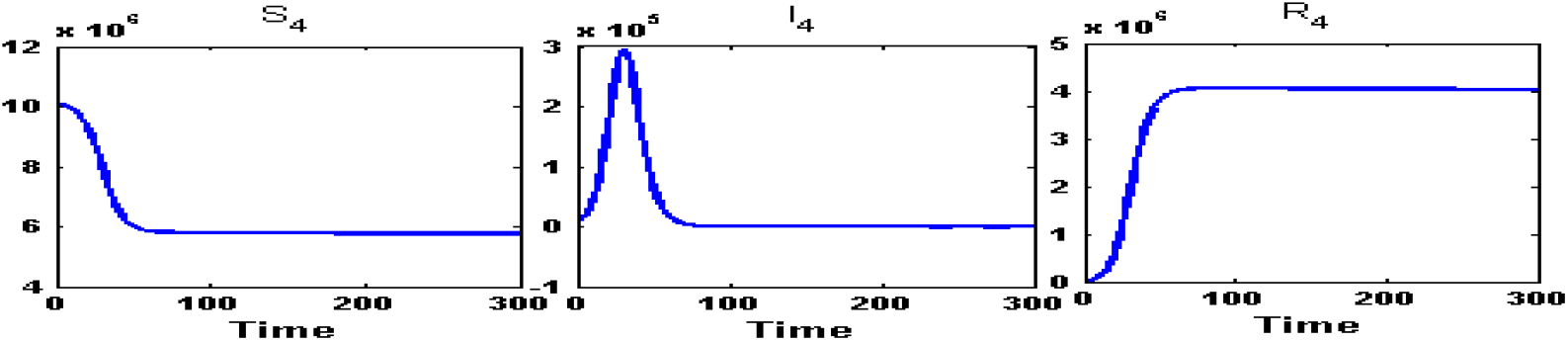
Susceptible (*S*_4_) class, Infected (*I*_4_) class and Recovered (*R*_4_) class people of North 24 Parganas(*n*_3_) for intra-district network. During the partial lock-down, the infected population will reach at its peak of an amount of 2,95,100 infected person in 31 days and will go below the initial infected in 66 days.

**Figure A.31:**
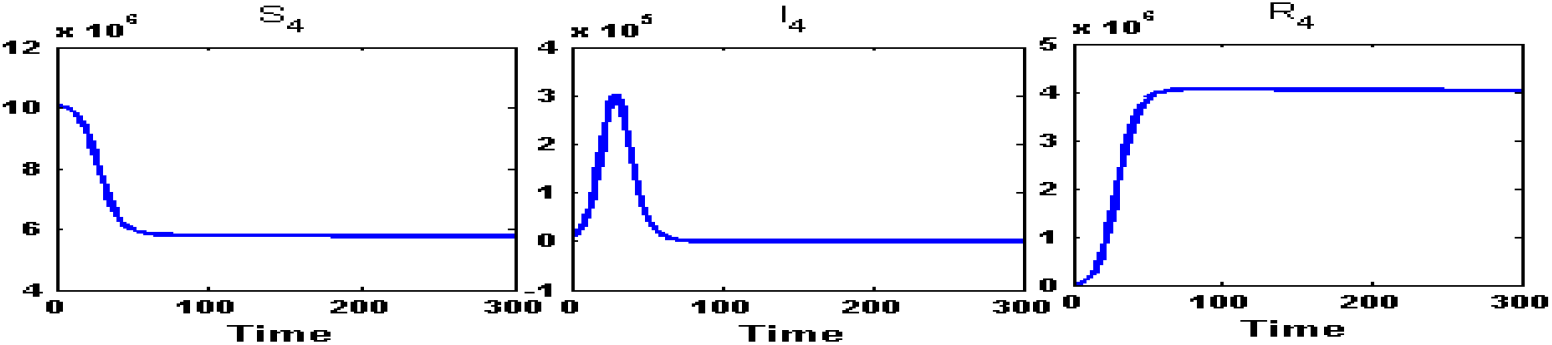
Susceptible (*S*_4_) class, Infected (*I*_4_) class and Recovered (*R*_4_) class people of North 24 Parganas(*n*_3_) for fully connected network. After the unlocking, within 30 days, the infected population will reach to its peak value at an amount 3,00,900 persons and down the initial amount of infection in 64 days.

**Figure A.32:**
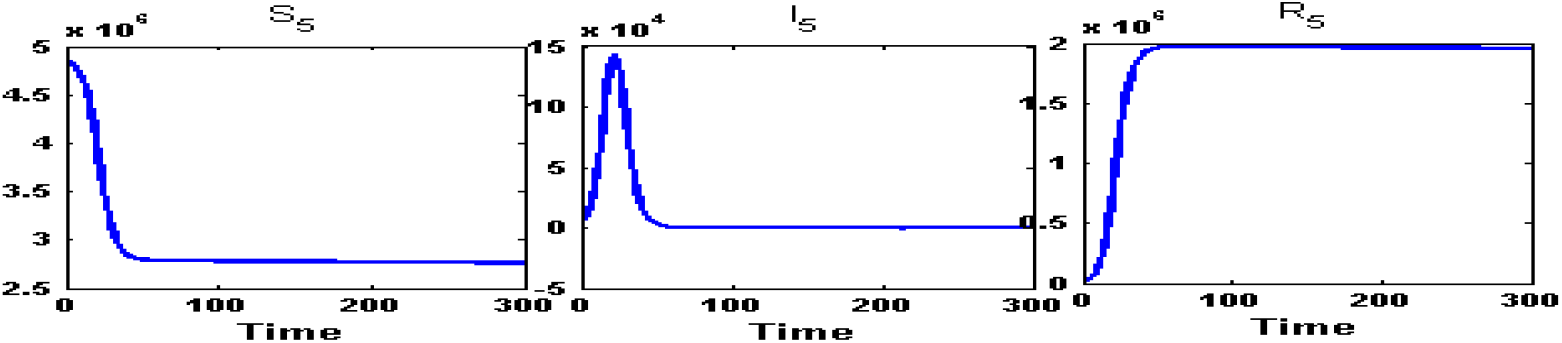
Susceptible (*S*_5_) class, Infected (*I*_5_) class and Recovered (*R*_5_) class people of Howrah(*n*_4_) for intra-district network. During the partial lock-down, the infected population will reach at its peak of an amount of 1,43,300 infected person in 22 days and will go below the initial infected in 48 days.

**Figure A.33:**
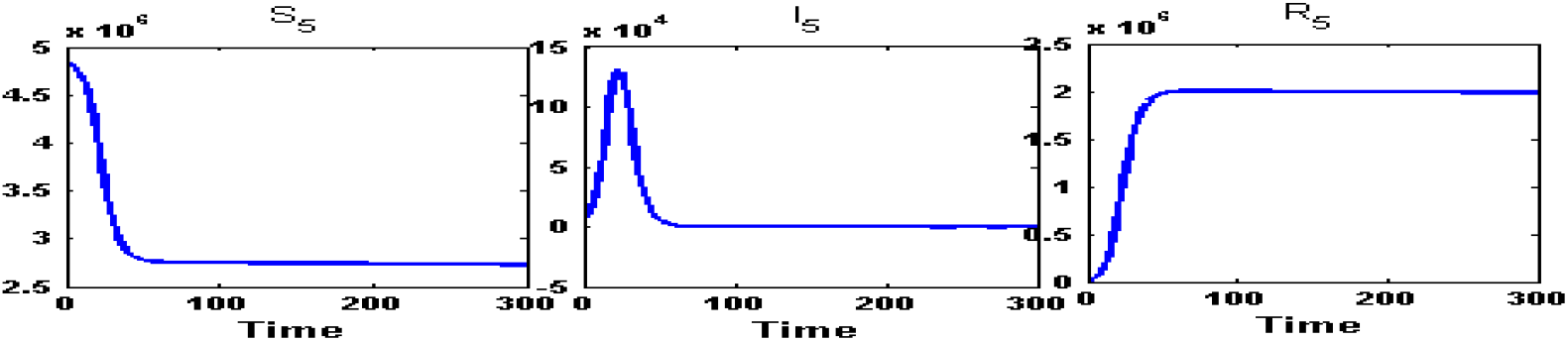
Susceptible (*S*_5_) class, Infected (*I*_5_) class and Recovered (*R*_5_) class people of Howrah(*n*_4_) for fully connected network. After the unlocking, within 53 days, the infected population will reach to its peak value at an amount 1,31,300 persons and down the initial amount of infection in 53 days.

**Figure A.34:**
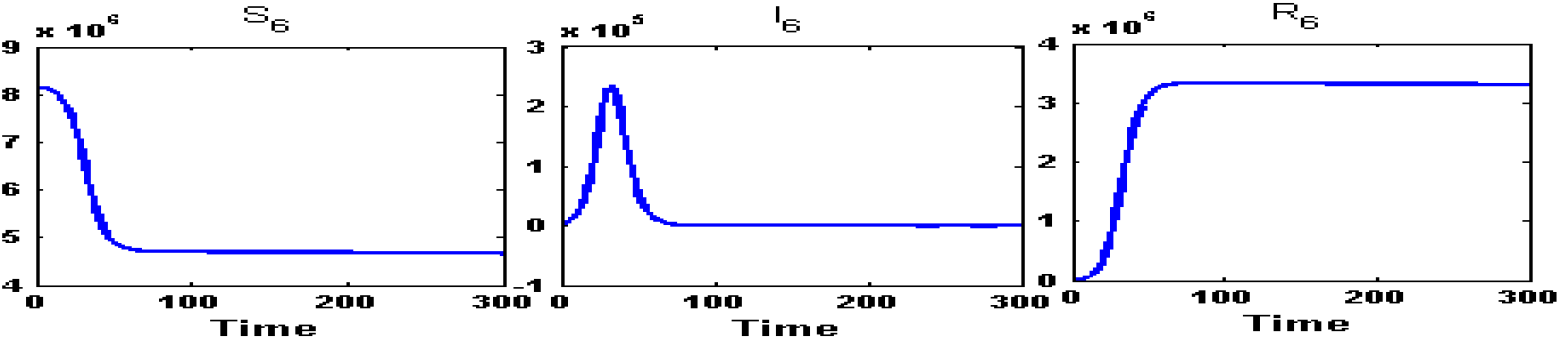
Susceptible (*S*_6_) class, Infected (*I*_6_) class and Recovered (*R*_6_) class people of South 24 Parganas(*n*_5_) for intra-district network. During the partial lock-down, the infected population will reach at its peak of an amount of 2,34,600 infected person in 33 days and will go below the initial infected in 72 days.

**Figure A.35:**
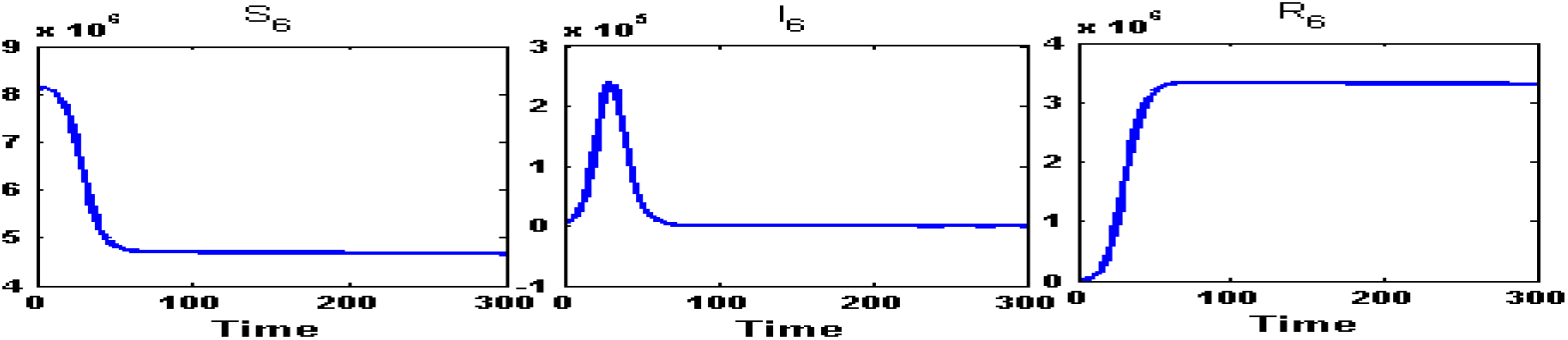
Susceptible (*S*_6_) class, Infected (*I*_6_) class and Recovered (*R*_6_) class people of South 24 Parganas(*n*_5_) for fully connected network. After the unlocking, within 30 days, the infected population will reach to its peak value at an amount 2,40,500 persons and down the initial amount of infection in 68 days.

**Figure A.36:**
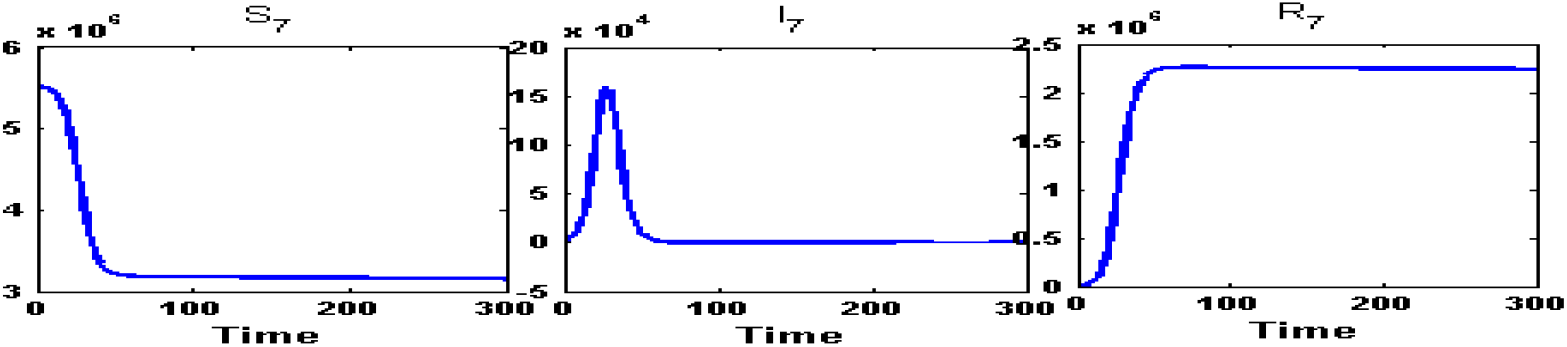
Susceptible (*S*_7_) class, Infected (*I*_7_) class and Recovered (*R*_7_) class people of Hooghly(*n*_6_) for intra-district network. During the partial lock-down, the infected population will reach at its peak of an amount of 1,60,000 infected person in 27 days and will go below the initial infected in 61 days.

**Figure A.37:**
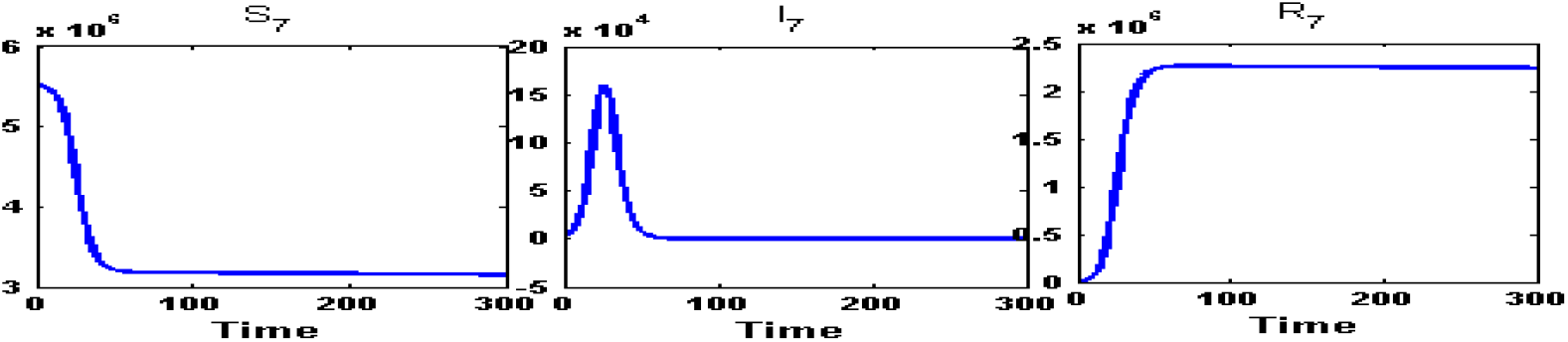
Susceptible (*S*_7_) class, Infected (*I*_7_) class and Recovered (*R*_7_) class people of Hooghly(*n*_6_) for fully connected network. After the unlocking, within 26 days, the infected population will reach to its peak value at an amount 1,59,700 persons and down the initial amount of infection in 59 days.

**Figure A.38:**
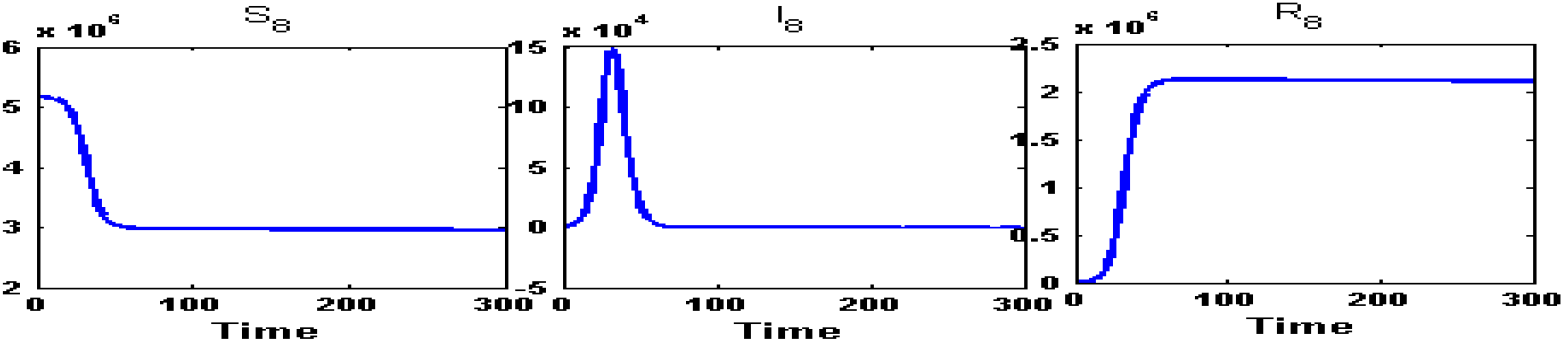
Susceptible (*S*_8_) class, Infected (*I*_8_) class and Recovered (*R*_8_) class people of Nadia(*n*_7_) for intra-district network. During the partial lock-down, the infected population will reach at its peak of an amount of 1,48,400 infected person in 33 days and will go below the initial infected in 70 days.

**Figure A.39:**
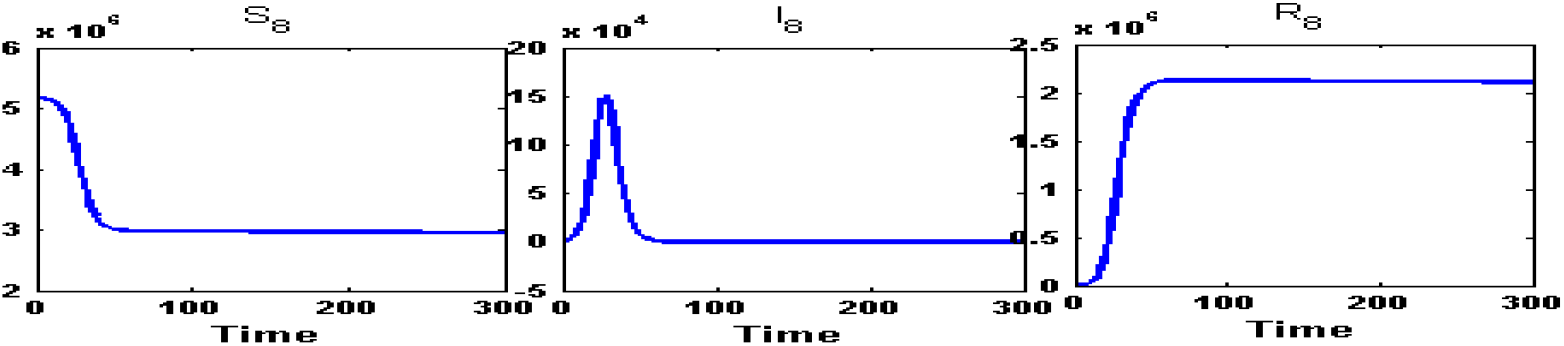
Susceptible (*S*_8_) class, Infected (*I*_8_) class and Recovered (*R*_8_) class people of Nadia(*n*_7_) for fully connected network. After the unlocking, within 28 days, the infected population will reach to its peak value at an amount 1,51,300 persons and down the initial amount of infection in 66 days.

**Figure A.40:**
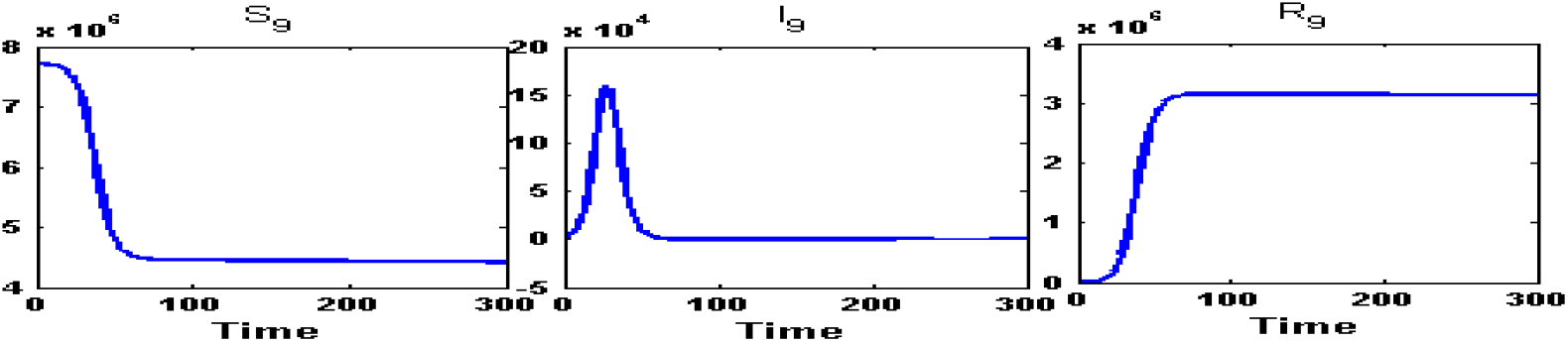
Susceptible (*S*_9_) class, Infected (*I*_9_) class and Recovered (*R*_9_) class people of Burdwan(*n*_8_) for intra-connected network. During the partial lock-down, the infected population will reach at its peak of an amount of 1,60,000 infected person in 27 days and will go below the initial infected in 66 days

**Figure A.41:**
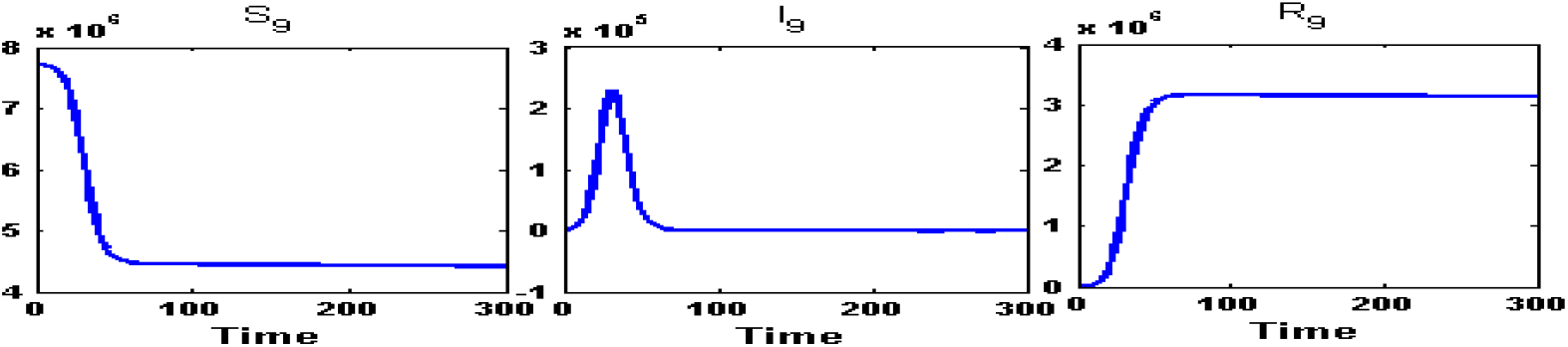
Susceptible (*S*_9_) class, Infected (*I*_9_) class and Recovered (*R*_9_) class people of Burdwan(*n*_8_) for fully connected network. After the unlocking, within 69 days, the infected population will reach to its peak value at an amount 2,28,800 persons and down the initial amount of infection in 76 days.

**Figure A.42:**
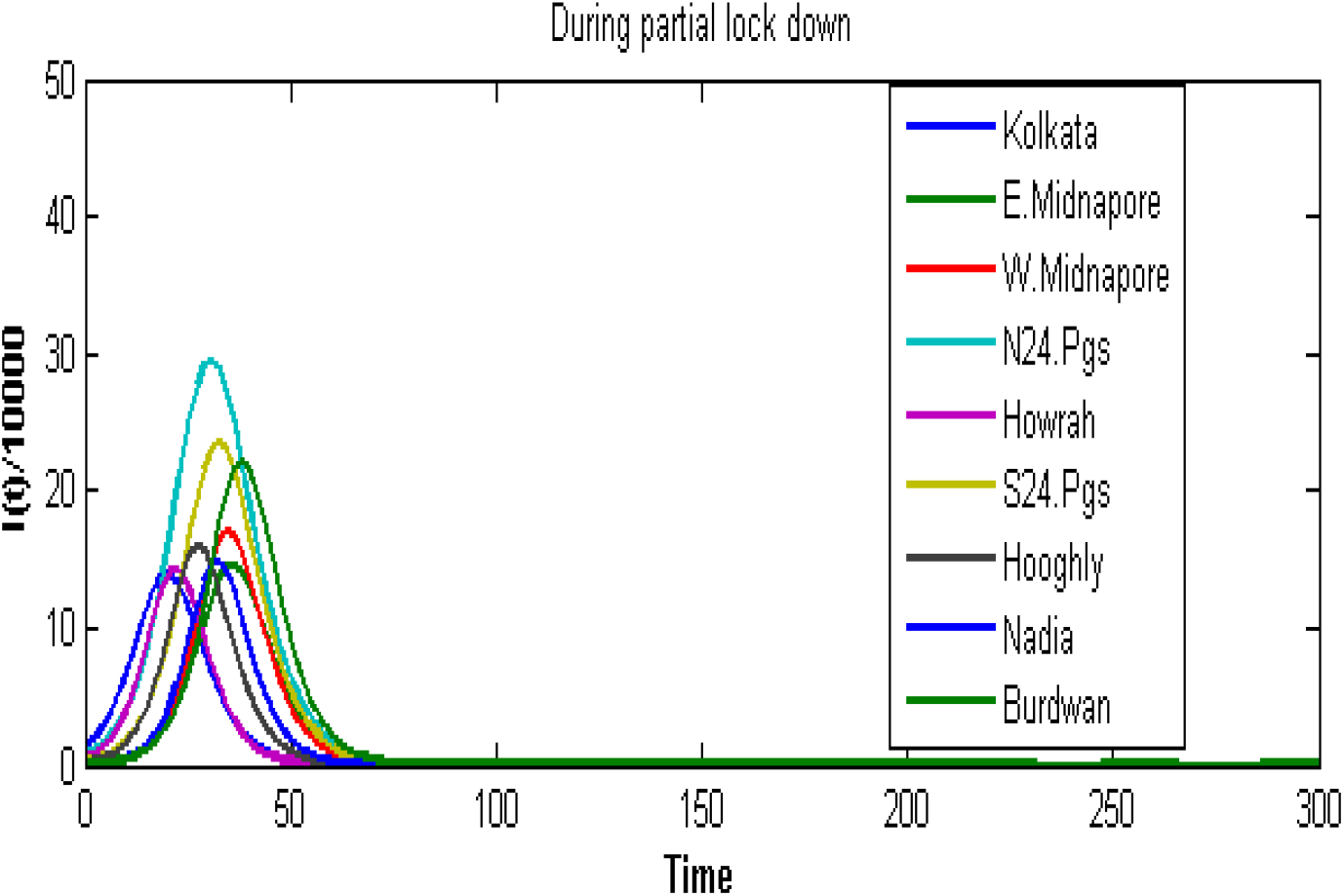
Infected population when the transportation open within their own network (Time Vs *I_i_/*10000). Infected population to reach at its peak for each of the nine districts will range between 21 to 33 days

**Figure A.43:**
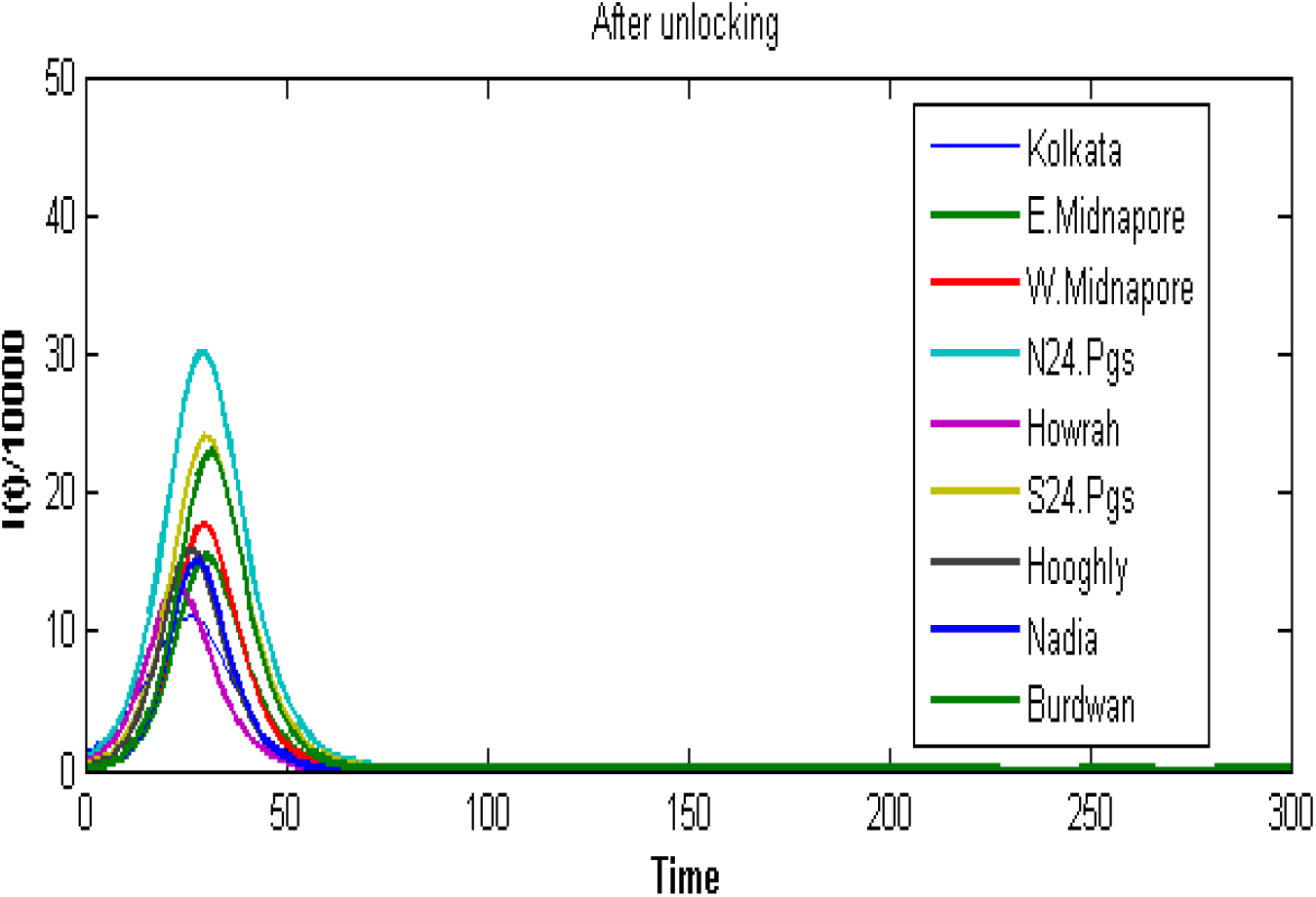
Infected population when the transportation open within the full network (Time Vs *I_i_/*10000). Infected population to reach at its peak for each of the nine districts will range between 23 to 32 days

## References

[1] T. P. Velavan, C. G. Meyer, The covid-19 epidemic, Tropical medicine & international health 25 (3) (2020) 278. 2

[2] Z. Wu, J. M. McGoogan, Characteristics of and important lessons from the coronavirus disease 2019 (COVID-19) outbreak in china: summary of a report of 72314 cases from the chinese center for disease control and prevention, Jama 323 (13) (2020) 1239–1242. 2

[3] W.-j. Guan, Z.-y. Ni, Y. Hu, W.-h. Liang, C.-q. Ou, J.-x. He, L. Liu, H. Shan, C.-l. Lei, D. S. Hui, et al., Clinical characteristics of coronavirus disease 2019 in China, New England journal of medicine 382 (18) (2020) 1708–1720. 2

[4] World health organization and others, coronavirus disease 2019 (COVID19): situation reports (2020). 2

[5] World health organization and others, coronavirus disease 2019 (COVID19): situation report, 126 (2020). 2

[6] R. David, India confirms its first coronavirus case (2020). 2

[7] The telegraph, 40,000 indians quarantined after ‘super spreader’ ignores government advice (2020). 2

[8] news.abplive.com, punjab: 27 bus drivers conductors that returned from nanded test corona positive. retrieved 6 may 2020 (2020). 2

[9] The hindu, 1,225 nanded returnees test COVID-19 positive: government. retrieved 14 may 2020 (2020). 2

[10] Outlook, all ASI-protected monuments, central museums across india to be shut till march 31: Govt. 2

[11] The economic times, uddhav thackeray imposes curfew in entire maharashtra. 23 march 2020. 2

[12] India today, coronavirus outbreak: States impose lockdown in battle against COVID-19 — all you need to know.retrieved 23 march 2020. 2

[13] Mohfw, press release 24 may 2020 [internet]. [cited 2020 may 25]. 2

[14] N. Fernandes, Economic effects of coronavirus outbreak (COVID-19) on the world economy, Available at SSRN 3557504 (2020). 2

[15] M. S. Majumder, C. Rivers, E. Lofgren, D. Fisman, Estimation of mers-coronavirus reproductive number and case fatality rate for the spring 2014 saudi arabia outbreak: insights from publicly available data, PLoS currents 6 (2014). 3

[16] S. Zhao, Q. Lin, J. Ran, S. S. Musa, G. Yang, W. Wang, Y. Lou, D. Gao, L. Yang, D. He, et al., Preliminary estimation of the basic reproduction number of novel coronavirus (2019-ncov) in china, from 2019 to 2020: A data-driven analysis in the early phase of the outbreak, International journal of infectious diseases 92 (2020) 214–217. 3

[17] J. M. Read, J. R. Bridgen, D. A. Cummings, A. Ho, C. P. Jewell, Novel coronavirus 2019-ncov: early estimation of epidemiological parameters and epidemic predictions, MedRxiv (2020). 3

[18] Y. Wang, Y. Wang, Y. Chen, Q. Qin, Unique epidemiological and clinical features of the emerging 2019 novel coronavirus pneumonia (COVID19) implicate special control measures, Journal of medical virology 92 (6) (2020) 568–576. 3

[19] L. Zou, F. Ruan, M. Huang, L. Liang, H. Huang, Z. Hong, J. Yu, M. Kang, Y. Song, J. Xia, et al., SARS-COV-2 viral load in upper respiratory specimens of infected patients, New England Journal of Medicine 382 (12) (2020) 1177–1179. 3

[20] R. M. Anderson, R. M. May, Infectious diseases of humans: dynamics and control, Oxford university press, 1992. 3

[21] O. Diekmann, J. A. P. Heesterbeek, Mathematical epidemiology of infectious diseases: model building, analysis and interpretation, Vol. 5, John Wiley & Sons, 2000. 3, 13

[22] H. W. Hethcote, The mathematics of infectious diseases, SIAM review 42 (4) (2000) 599–653. 3

[23] F. Brauer, C. Castillo-Chavez, C. Castillo-Chavez, Mathematical models in population biology and epidemiology, Vol. 2, Springer, 2012. 3

[24] W. O. Kermack, A. G. McKendrick, A contribution to the mathematical theory of epidemics, Proceedings of the royal society of london. Series A, Containing papers of a mathematical and physical character 115 (772) (1927) 700–721. 3

[25] Q. Lin, S. Zhao, D. Gao, Y. Lou, S. Yang, S. S. Musa, M. H. Wang, Y. Cai, W. Wang, L. Yang, et al., A conceptual model for the outbreak of coronavirus disease 2019 (COVID-19) in wuhan, china with individual reaction and governmental action, International journal of infectious diseases 93 (2020) 211–216. 3

[26] C. Anastassopoulou, L. Russo, A. Tsakris, C. Siettos, Data-based analysis, modelling and forecasting of the COVID-19 outbreak, PloS one 15 (3) (2020) e0230405. 3

[27] F. Casella, Can the COVID-19 epidemic be managed on the basis of daily data?, arXiv preprint arXiv:2003.06967 (2020). 3

[28] J. T. Wu, K. Leung, M. Bushman, N. Kishore, R. Niehus, P. M. de Salazar, B. J. Cowling, M. Lipsitch, G. M. Leung, Estimating clinical severity of COVID-19 from the transmission dynamics in Wuhan, China, Nature Medicine 26 (4) (2020) 506–510. 3

[29] N. Wang, Y. Fu, H. Zhang, H. Shi, An evaluation of mathematical models for the outbreak of COVID-19, Precision Clinical Medicine 3 (2) (2020) 85–93. 3

[30] Y. Fang, Y. Nie, M. Penny, Transmission dynamics of the COVID-19 outbreak and effectiveness of government interventions: A data-driven analysis, Journal of medical virology 92 (2020) 645–659. 3

[31] C. Hou, J. Chen, Y. Zhou, L. Hua, J. Yuan, S. He, Y. Guo, S. Zhang, Q. Jia, C. Zhao, et al., The effectiveness of the quarantine of Wuhan city against the corona virus disease 2019 (COVID-19): well-mixed SEIR model analysis, Journal of Medical Virology 92 (2020) 841–842. 3

[32] A. Radulescu, K. Cavanagh, Management strategies in a SEIR model of COVID 19 community spread, arXiv preprint arXiv:2003.11150 (2020). 4

[33] G. Pandey, P. Chaudhary, R. Gupta, S. Pal, SEIR and regression model based COVID-19 outbreak predictions in India, arXiv preprint arXiv:2004.00958 (2020). 4

[34] J. Hellewell, S. Abbott, A. Gimma, N. I. Bosse, C. I. Jarvis, T. W. Russell, J. D. Munday, A. J. Kucharski, W. J. Edmunds, F. Sun, et al., Feasibility of controlling COVID-19 outbreaks by isolation of cases and contacts, Lancet Global Health 8 (4) (2020) 488–496. 4

[35] A. J. Kucharski, T. W. Russell, C. Diamond, Y. Liu, J. Edmunds, S. Funk, R. M. Eggo, F. Sun, M. Jit, J. D. Munday, et al., Early dynamics of transmission and control of COVID-19: a mathematical modelling study, The lancet infectious diseases (2020). 4

[36] F. Ndairou, I. Area, J. J. Nieto, D. F. Torres, Mathematical modeling of COVID-19 transmission dynamics with a case study of wuhan, Chaos, Solitons & Fractals (2020) 109846. 4

[37] D. Khatua, A. De, S. Kar, E. Samanta, S. M. Mandal, A dynamic optimal control model for SARS-COV-2 in India, Available at SSRN 3597498 (2020). 4

[38] D. Khatua, A. De, S. Kar, E. Samanta, A. A. Seikh, D. Guha, A fuzzy dynamic optimal model forCOVID-19 epidemic in India based on granular differentiability, Available at SSRN 3621640 (2020). 4

[39] S. Boatto, L. M. Stolerman, J. Pacheco, F. Santos, R. S. Khouri, SIR-network model for epidemics dynamics in a city, preprint (2014). 4, 5

[40] Y. Hu, L. Min, Y. Su, Y. Kuang, Mathematical analysis of an SIR network model with imperfect vaccination and varying size of population, in: Proceedings of the 8th International Conference on Computer Modeling and Simulation, 2017, pp. 7–13. 4

[41] F. Ball, T. Britton, K. Y. Leung, D. Sirl, A stochastic SIR network epidemic model with preventive dropping of edges, Journal of mathematical biology 78 (6) (2019) 1875–1951. 4

[42] L. M. Stolerman, D. Coombs, S. Boatto, SIR-network model and its application to dengue fever, SIAM Journal on Applied Mathematics 75 (6) (2015) 2581–2609. 4, 8, 13

[43] C. Atkinson, G. Reuter, Deterministic epidemic waves, in: Mathematical Proceedings of the Cambridge Philosophical Society, Vol. 80, Cambridge University Press, 1976, pp. 315–330. 4

[44] G. Fulford, M. Roberts, J. Heesterbeek, The metapopulation dynamics of an infectious disease: tuberculosis in possums, Theoretical population biology 61 (1) (2002) 15–29. 4

[45] J. Arino, P. Van den Driessche, A multi-city epidemic model, Mathematical Population Studies 10 (3) (2003) 175–193. 4, 6

[46] M. J. Keeling, L. Danon, M. C. Vernon, T. A. House, Individual identity and movement networks for disease metapopulations, Proceedings of the National Academy of Sciences 107 (19) (2010) 8866–8870. 4

[47] P. Van den Driessche, J. Watmough, Reproduction numbers and subthreshold endemic equilibria for compartmental models of disease transmission, Mathematical biosciences 180 (1-2) (2002) 29–48. 7, 13

[48] O. Diekmann, J. A. P. Heesterbeek, J. A. Metz, On the definition and the computation of the basic reproduction ratio \_0_ in models for infectious diseases in heterogeneous populations, Journal of mathematical biology 28 (4) (1990) 365–382. 8, 13

[49] List of districts of West Bengal from: https://en.wikipedia.org/wiki/list of districts of West Bengal (2017). 9

[50] Department of health and family welfare,West Bengal, Corona Bulletin 1 (2020). 9

[51] Covid-19 pandemic in West Bengal, retrieved may. 04, 2020 from: en.wikipedia.org/wiki/covid-19 pandemic in West Bengal (2020). 10

[52] Encyclopaedia britannica, Encyclopaedia Britannica, Incorporated, 1957. 11

[53] T. Dey, Suburban railway network of Kolkata: A geographical appraisal (2012). 11

[54] Age structure and marital status, census 2011 available from: http://censusindia.gov. in, Census And You/age struct ure and maritxal status. aspx (Accessed on 12.11. 2017). 12

[55] Indiaspend, how many people does one COVID-19 patient infect in india? retrieved 29 may, 2020 (2020). 13

